# MET receptor activation by stromal cells serves as promising target in melanoma brain metastases

**DOI:** 10.1101/2023.10.16.23297080

**Authors:** Torben Redmer, Elisa Schumann, Kristin Peters, Martin E. Weidemeier, Stephan Nowak, Henry W.S. Schroeder, Anna Vidal, Helena Radbruch, Annika Lehmann, Susanne Kreuzer-Redmer, Karsten Jürchott, Josefine Radke

## Abstract

The development of brain metastases hallmarks disease progression in 20-40% of melanoma patients and is a serious obstacle to therapy. Understanding the processes involved in the development and maintenance of melanoma brain metastases (MBM) is critical for the discovery of novel therapeutic strategies. Here, we generated transcriptome and methylome profiles of MBM showing high or low abundance of infiltrated Iba1^high^ tumor-associated microglia and macrophages (TAMs). Our survey identified potential prognostic markers of favorable disease course and response to immune checkpoint inhibitor (ICi) therapy, among them *APBB1IP* and the interferon-responsive gene *ITGB7*. In MBM with high ITGB7/APBB1IP levels, the accumulation of TAMs correlated significantly with the immune score. Signature-based deconvolution of MBM via single sample GSEA revealed enrichment of interferon-response and immune signatures and revealed inflammation, stress and MET receptor signaling. MET receptor phosphorylation/activation maybe elicited by inflammatory processes in brain metastatic melanoma cells via stroma cell-released HGF. We observed phospho-MET^Y1234/1235^ in a subset of MBM and observed marked response of brain metastasis-derived cell lines (BMCs) that lacked druggable BRAF mutations or developed resistance to BRAF inhibitors (BRAFi) *in vivo* to MET inhibitors PHA-665752 and ARQ197 (tivantinib). In summary, the activation of MET receptor in brain colonizing melanoma cells by stromal cell-released HGF may promote tumor cells self-maintenance and expansion might counteract ICi therapy. Therefore, therapeutic targeting of MET possibly serves as promising strategy to control intracranial progressive disease and improve patient survival.

## Introduction

The interaction of brain colonizing tumor cells with the tumor microenvironment (TME), mainly comprising innate and adaptive immune cells, microglia, astrocytes, neurons and oligodendrocytes crucially determines the developmental stages of brain metastases (BM). Brain metastases are observed in 20 – 40% of melanoma patients during the course of disease and micrometastases are evident in more than 75% of autopsied brains^1^. Hence, only a subset of melanoma cells that entered the brain develop symptomatic and detectable BM during the lifetime of melanoma patients. Unlike peripheral metastases, the emergence of BM depends on a plethora of environmental cues such as the spatiotemporal availability of factors that are provided by cells of the TME, supporting or repressing tumor cell growth^2^. Moreover, single-cell RNA sequencing (scRNAseq) studies have confirmed regional heterogeneity of astrocytes^3^, oligodendrocytes^4^ and microglia^5,6^ in healthy human brains. Particularly, astrocytes and microglia adopt a reactive cell state^7,8^ that accompanies secretion of pro- and anti-inflammatory factors under pathological conditions^9, 10^. It is therefore possible that subfractions of astrocytes and microglia communicate and react with tumor cells in different ways. Probably, neuroinflammation precedes colonization of the brain by tumor cells. However, tumor cells invading the brain amplify inflammatory processes mediated by astrocytes and infiltrating tumor-associated microglia and macrophages (TAMs)^11,12^. Recently, signaling mediated by hepatocyte growth factor (HGF) and the related receptor MET (c-MET, HGFR) was identified as the trigger of reactive microglia^13^. HGF is thereby secreted by microglia in the context of trauma but also under normal conditions and seems to play a special role in the growth and self-renewal of neural stem cells in the subventricular zone (SVZ) of rat brains^14,15^. Therefore, metastatic melanoma cells expressing MET might scavenge HGF from the brain for activation of processes downstream of MET mediating survival and proliferation.

Here, we used transcriptome and methylome profiling to unravel the epigenetic and transcriptomic landscapes of MBM that featured infiltration of TAMs with emphasis on the potential role of microglia in the activation of the HGF/MET receptor signaling pathway. The MET receptor inhibitors PHA-665752 and tivantinib (ARQ197) effectively blocked the growth of brain metastases derived cells (BMCs). Hence, targeting MET receptor signaling might serve as a potent therapeutic target for brain metastases lacking druggable BRAF^V600^ mutations.

## Results

### A microglia-specific gene cluster discriminates MBM

Microglia are a unique population of antigen-presenting cells in the central nervous system (CNS) that are capable of clearing the brain of microbes, dead cells and protein aggregates^16^. Besides, microglia play a crucial role during injury repair and display an exceptional role in immune surveillance and tumor clearance^17,18^. Although the role of tumor-associated microglia and macrophages (TAMs) in primary brain tumors such as glioblastoma^19–22^ has been intensively studied, their role in the progression of brain metastases remains poorly understood.

We performed immunohistochemistry (IHC) of our MBM cohort (Supplementary table 1 and^23^) to determine the levels of activated, Iba1^high^ TAMs. Although Iba1/AIF1 serves as a well-established marker, reactive microglia cannot be distinguished from brain infiltrated macrophages^24^. Initial studies of MBM revealed that Iba1/AIF1 levels classified tumors into highly and lowly TAM infiltrated (Figure 1a, Supplementary figure 1a). Moreover, we observed overlapping patterns of infiltration of Iba1^high^ TAMs and CD3^+^ T cells (Figure 1b). As CD3 only provided information about levels of T cell infiltration, we used the ESTIMATE algorithm^25^ to gain insight into the overall degree of immune cell infiltration of MBM. In line with our previous observation, tumors with intensive TAM and T cell infiltration exhibited a high immune score (Pts 3, 4, 10, 12) whereas MBM with low levels of Iba1^high^/ CD3^+^ cell infiltration (Pts 1, 2) or low expression of Iba1/AIF1 (Supplementary figures 1b, c) showed low immune scores (Figure 1c). As expected, brain metastases derived cell lines (BMCs) with absence of immune cells featured lowest scores (Supplementary figures 1b, c). The brain has long been considered a sanctuary where tumor cells can grow undisturbed and protected from attack by immune cells. Therefore, we next investigated expression levels of Iba1/AIF1 in brain (MBM) and extracranial metastases (EM). We observed AIF1 expression in both as well as a high correlation with immune score (Figure 1d), suggesting a relationship between levels of infiltration of TAMs and immune cells not only in the brain. As high levels of immune cell/T cell infiltration are generally associated with good prognosis^26^, we determined the probability of survival related to Iba1/AIF1 expression of patient’s with (study EGAS00001003672) and without (TCGA-SKCM) MBM. We observed beneficial effects of high Iba1/AIF1 levels in the TCGA cohort (HR=0.46 (0.35 – 0.62), logrank p=1.3e-07) (Figures 1e, f), however, Iba1/AIF1 level had no beneficial effects on the survival of MBM patients. Since no data on TAM-infiltrated MBM are available, we performed comparative methylome and transcriptome profiling of Iba1^high^ (n=5-10) and Iba1^low^ (n=2-6) tumors and identified a set of 417 differentially methylated genomic regions (DMRs) that corresponded to 294 MBM expressed genes (Figure 1g) a core set of markers (n=31) sufficient to split tumors (Figure 1h; Supplementary tables 2, 3). Among them, we identified the integrin family member and gut-homing receptor ITGB7 -which we described in our previous study as a distinguishing mark between BRAF and NRAS mutant MBM^23^ - and *APBB1IP* (amyloid b precursor protein-binding family b member 1 interacting protein). Both are associated with better prognosis in patients with colorectal cancer^27,28^ and clustered with known TAM-associated genes such as *P2RY12* and *AIF1* (Figure 1h). Remarkably, all clustered tumors were associated with a high immune score. A correlation analysis of clustered genes revealed a high degree of correlation among each other (Figure 1i) and association with hepatocyte growth factor (HGF) that was recently connected with microglia activation^13^. However, only some of the identified markers within the gene cluster were specifically expressed in microglia but not in brain infiltrating macrophages or other brain cells such as *APBB1IP* (Figure 1j). The latter gene which has been identified as a conserved microglial gene^29^ and binding partner of amyloid precursor protein (APP), Tau, 14-3-3g, and glycogen synthase kinase 3 b (GSK3 b) was associated with actin dynamics and retinoic acid signaling^30,31^. Expression of APBB1IP was significantly (MBM: R=0.86, p<2.2e-16) correlated with immune score (Figure 1k) and survival of melanoma patients (Supplementary figures 1d-e). Moreover, our survey identified a differentially methylated side (Supplementary table 4) within the promoter of PD-L2 (PDCD1LG2) that may predict progression-free survival in melanoma patients receiving anti-PD-1 immunotherapy^32^. PD-L2 expression was associated with favorable survival (p=0.020) of patients with MBM (Supplementary figure 1f). We found additional genes among our cluster that were expressed in TAMs and significantly associated with immune score (Supplementary figures 1g-n).

**Figure 1:**
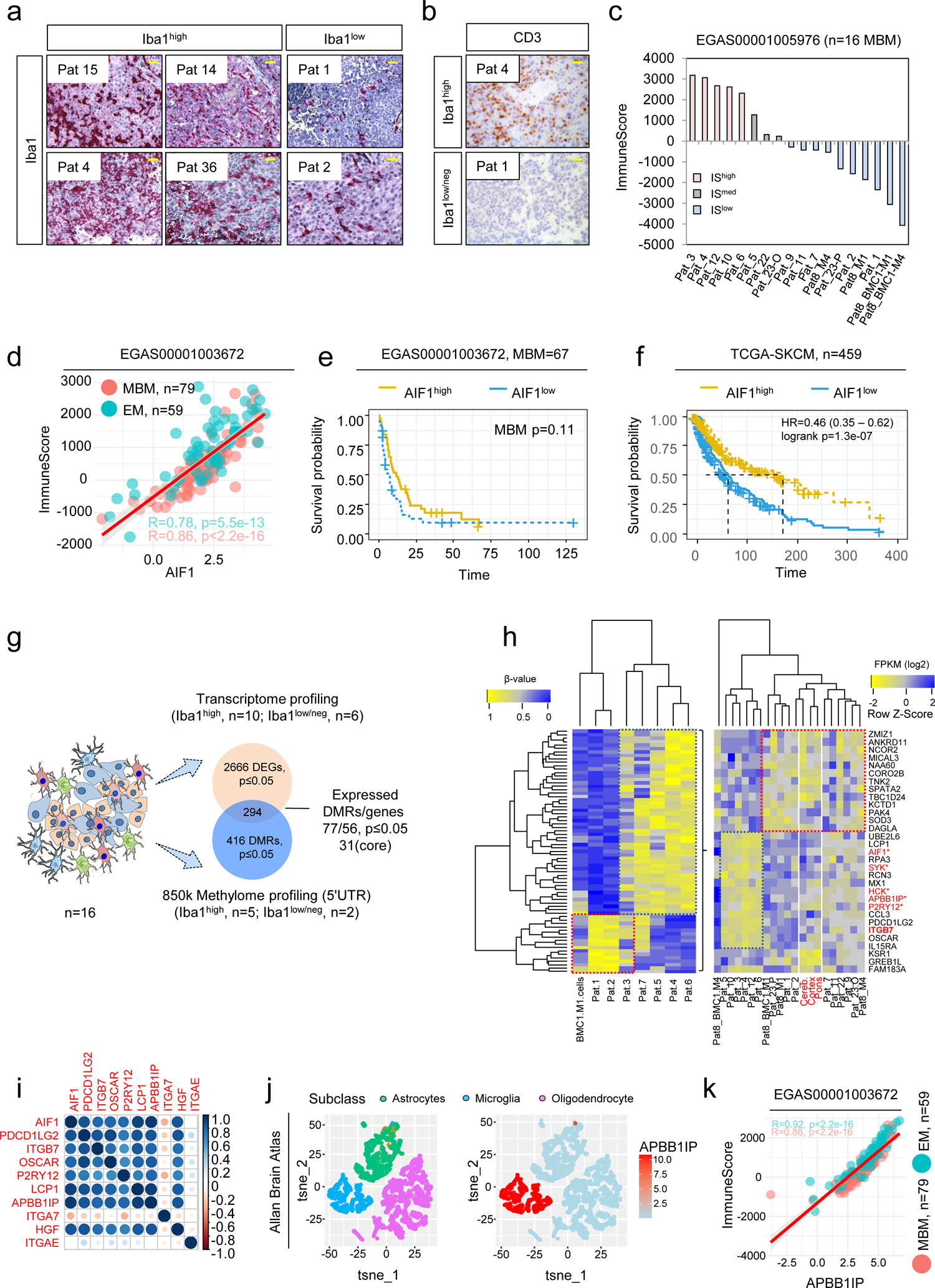
Transcriptome and methylome profiling of Iba1^high^ and Iba1^neg^ MBM revealed identification of subset-specific genes. a.) Immunohistochemistry (IHC) for Iba1 (red) of MBM of indicated patients. b.) Representative IHC for levels of CD3 in Iba1^high^ (Pat 4) and Iba1^neg^ (Pat 1) MBM. c.) Immune score of MBM (study EGAS00001005976, n=16) indicating different immunologic (color coded) subsets of tumors. d.) Dot plot showing the significant correlation of Iba1/AIF1 expression and immune score of brain metastases (BM, R=0.86, p<2.2e-16) and extracranial metastases (EM, R=0.78, p=5.5e-13). e.) Survival analysis of patients with MBM (study, EGAS00001003672), featuring high or low level of Iba1/AIF1 expression revealed no significant difference (p=0.11). f.) Survival analysis of TCGA melanoma patients (n=459), featuring high or low level of Iba1/AIF1 expression revealed significant difference (logrank p=1.3e-07) and Cox-regression analysis showed association with favorable disease course (HR=0.46). g.) Schematic representation of candidate identification by methylome and transcriptome profiling of n=16 MBM of study. Methylome (850k) profiling of Iba1^high^ (n=5) or Iba1^neg^ (n=2) identified 416 differentially methylated regions (DMRs), within the 5’-UTR of 316 corresponding genes of which 296 were expressed in MBM with 56 genes (77 DMRs), significantly (p≤0.05) discriminating Iba1^high^ and Iba1^low/neg^ MBM. h.) Heat map representation of 77 DMRs (left panel) and top expressed (right panel) genes (n=31). Analysis identified a panel of 12 genes that clustered with expression of microglia/TAM-associated genes AIF1, SYK and HCK. i.) Correlation analysis of cluster genes with association to immune/TAM regulated processes, strength of correlation is color coded. j.) Comparative t-SNE representation of brain cell subclasses microglia, neurons and oligodendrocytes (left) and expression of *APBB1IP* (Amyloid Beta Precursor Protein Binding Family B Member 1 Interacting Protein), expression level (log2 RPKM) is color coded. k.) Dot plot showing the significant correlation of *APBB1IP* expression and immune score of brain metastases (BM, R=0.86, p<2.2e-16) and extracranial metastases (EM, R=0.92, p<2.2e-16). Significance was determined by unpaired, two-sided t-test (d, g, k).

### Expression of ITGB7 serves as indicator of immune cell infiltration

Recent studies have shown that ITGB7 plays a critical role in the recruitment of T cells to the intestine and that downregulation of ITGB7 is important in protecting intestinal tumors from attack by activated T cells^27,33^. Hence, we sought to investigate *ITGB7* in more detail. Mining of publicly available immune cell data (studies GSE146771^34^, DICE database^35^) revealed expression of *ITGB7* across different immune cell stages including naïve and memory subsets of T cells, B cells and NK cells (Figure 2a and Supplementary figure 2a). We found that *ITGB7* was rather expressed in MBM with infiltration of immune cells and particularly within immune cell dense areas (Supplementary figure 2b). Co-staining revealed accumulation of CD3^+^ T cells as well as of Iba1^high^ TAMs (Figure 2b). Ranking of MBM regarding levels of *ITGB7* expression showed co-occurrence in the expression of CD4, CD274, Sushi Domain Containing 3 (SUSD3) and *ITGB7* level (Figure 2c) and validated a possible, previously observed^23^ correlation of *ITGB7* and SUSD3. Moreover *ITGB7, SUSD3* and *APBB1IP* showed expression across different immune cell types except for monocytes and NK cells (Supplementary figures 2c-f). Global (850k) methylome profiling uncovered four epigenetic regulation sites of *ITGB7* (Supplementary table 4) with two sites that were associated with expression levels and immune score (Figures 2d, left and center panel, Supplementary figure 3a), located in a proximal enhancer-like region (probe cg26689077) or near by the promotor of ITGB7 (probe cg01033299). The latter site was also identified in the TCGA-SKCM cohort. The sites did not correlate with the BRAF mutation status of MBM (Figures 2d, right panel) in contrast to additional two sides that were found within intergenic regions including an CpG island located between exons 4 and 5 (probes cg11510999 and cg18320160; Supplementary figures 3b-e). Hence, methylome profiling of MBM identified two DMRs within the *ITGB7* gene that might serve as indicators of the degree of immune cell infiltration.

**Figure 2:**
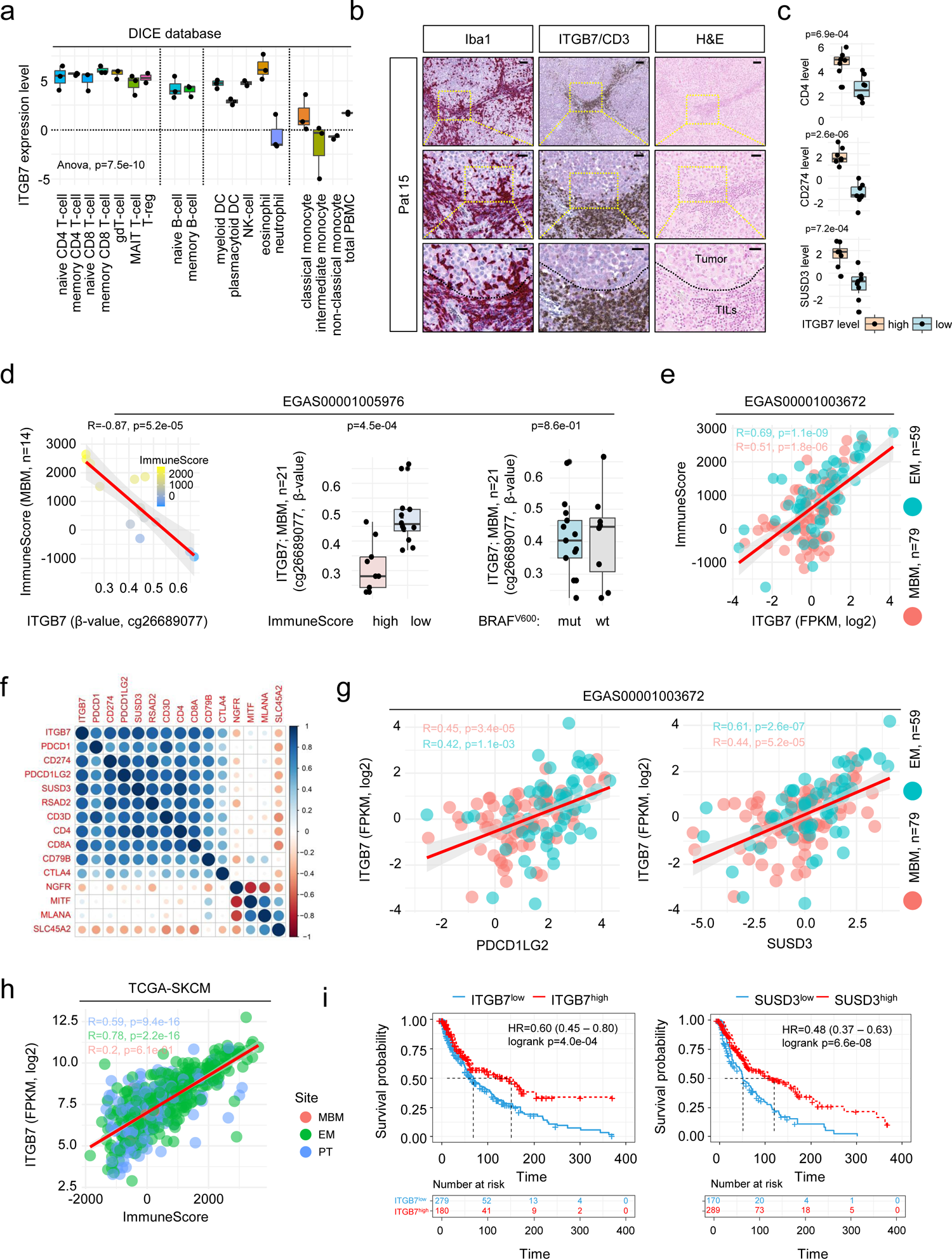
Expression of ITGB7 serves as indicator of lymphocyte infiltration. a.) Box plot representation of levels of ITGB7 indicates a wide pattern of expression among indicated immune cell populations. Monocytes and neutrophil granulocytes show low levels of ITGB7. b.) IHC of a representative MBM of a patient with refractory intracranial disease for Iba1 (red, first column) and CD3 (brown, second column) indicating focal enrichment of microglia/macrophages and CD3^+^ T cells within ITGB7 positive areas (red, second column). Hematoxylin and eosin (H&E) staining shows discrimination of tumor cells and tumor-infiltrating lymphocytes (TILs) c.) Expression (FPKM, log2) of CD4, PD-L1 (CD274) and SUSD3 in MBM with high or low level of ITGB7, indicating cellular co-occurrence. d.) Dot plot showing the significant inverse correlation (R=-0.87, p=5.2e-05) of β-values (probe cg26689077) indicating the methylation level at a side located within the proximal enhancer-like structure of the ITGB7 gene and immune score of MBM (n=14) of study EGAS00001005976 (first panel). Box plots represent a significant (p=4.5e-04) or non-significant (p=0.86) association of ITGB7 methylation (probe cg26689077) or BRAF mutation status (center and right panels) of all MBM investigated (n=21). e.) Dot plot showing the significant correlation of ITGB7 expression and immune score of MBM (R=0.51, p=1.8e-06) and EM (R=0.61, p=1.1e-09) indicating immune-related expression of ITGB7 irrespective of the side of metastasis. f.) Correlation map showing high association (p<0.05) of ITGB7 with relevant immune cell markers such as PD-1 (PDCD1), PD-L1 (CD274), PD-L2 (PDCD1LG2) but low correlation with tumor cell markers NGFR, MITF, MLANA or SLC45A2. g.) Dot plot showing the significant correlation of ITGB7 and expression of PD-L2 (BM: R=0.45, p=3.4e-05; EM: R=0.42, p=1.1e-03) and SUSD3 (BM: R=0.44, p=5.2e-05; EM: R=0.61, p=2.6e-07). h.) Dot plot showing the correlation of ITGB7 expression and immune score of primary (PT; R=0.59, p=9.4e-16), metastatic (EM; R=0.78, p=2.2e-16) and brain metastatic (BM; R=0.2, p=0.61) melanoma (TCGA-SKCM), indicating that expression of ITGB7 is independent from melanoma progression stages. i.) Survival analysis of TCGA melanoma patients (n=459), featuring high or low level of ITGB7 and SUSD3 expression revealed significant difference (logrank p=4.0e-04 and p=6.6e-08) and Cox-regression analysis showed association with favorable disease course (HR=0.60 and HR=0.48). Box and whisker plots show median (center line), the upper and lower quartiles (the box), and the range of the data (the whiskers), including outliers (a, c, d). Significance was determined by unpaired, two-sided t-test (c, d) or one-way ANOVA (a).

A recent study demonstrated that MBM feature a lower T cell content than matched extracranial metastases, however response rates to ICi of both were comparable^36^. Assuming that ITGB7 expression might be crucial for T cell recruitment, we ascertained *ITGB7* levels in MBM (n=79) and EM (n=59; study EGAS00001003672). We observed that *ITGB7* was expressed in both metastatic subtypes and was significantly correlated (MBM: R=0.51, p=1.8e-06; EM: R=0.69, p=1.1e-09) with the tumoŕs immune scores (Figure 2e). As we suggest that *ITGB7* expression might indicate the degree of immune cell infiltration and possibly serve as indicator of response to ICi, we next performed correlation analysis of *ITGB7* and known markers of T cells and B cells. We observed a high concordance with immune cell-related but not tumor cell-related genes (*NGFR, MITF*, *MLANA, SLC45A2*) and correlation with expression of *PDCD1LG2* (PD-L2) and *SUSD3*, irrespective of the side of metastasis (Figures 2f, g). In line with previous observations, *ITGB7* was expressed in primary and metastatic tumors (TCGA-SKCM) and like SUSD3 was associated with favored survival (Figure 2i). In summary, our survey identified a set of markers that are potentially associated with the level of TAM/immune cell infiltration, particularly *ITGB7* might serve as a marker for a favorable course of the disease.

### A signature-based deconvolution revealed MET receptor signaling in microglia-enriched MBM

Our previous survey identified a set of markers that potentially characterize a molecular subset of MBM, likely showing a favorable course and response to ICi therapy^37,38^. To further characterize this set of tumors, we performed single-sample Gene set Enrichment-Analysis (ssGSEA) using defined immune-related and gene signatures specifying signaling processes such as MET receptor or STAT3 signaling among others that are reported to be involved in immune-response mechanisms (Supplementary table 5). We observed that MBM featuring a high immune score showed activation of MET and STAT3 signaling, increased tumor inflammation, stress and senescence (SenMayo^39^) (Figure 3a). Moreover, deconvolution revealed the presence of reactive microglia, astrocytes and immune cell subsets, among them stem cell-like CD8^+^ T cells (TCF7)^40^ in tumors, absent in BMCs. CD8^+^ (TCF7) T cells are necessary for long-term maintenance of T cell responses and predicted positive clinical outcome^41,42^. Signatures clearly discriminated MBM and BMCs and reinforced the differences of Iba1^high^ (Pts 3, 4) and Iba1^low/neg^ (Pts 1, 2) tumors. We therefore suggest that the activation of MET- or STAT3-mediated signaling processes or those related to stress/senescence or inflammation strongly depend on the composition of the tumor microenvironment, likely determining the response to therapeutic interventions. Although infiltration of TAMs is not evident in all MBM, microglia infiltration seems to be an early occurring process observed ∼21d after intracranial injection of BMCs into brains of immune compromised Crl:CD1-Foxn1^nu^ mice^23^ (Figure 3b). Moreover, we observed activation of Stat3 signaling in tumor adjacent cells (Figure 3b), suggesting that brain microenvironmental cells are activated after a short time of tumor-stroma interaction and establish an inflammatory environment. We performed ssGSEA and applied the above mentioned signatures and observed a comparable pattern of enrichment in a more comprehensive and independent set of MBM (study EGAS00001003672^43^, n=79 MBM) (Supplementary figure 4a).

**Figure 3:**
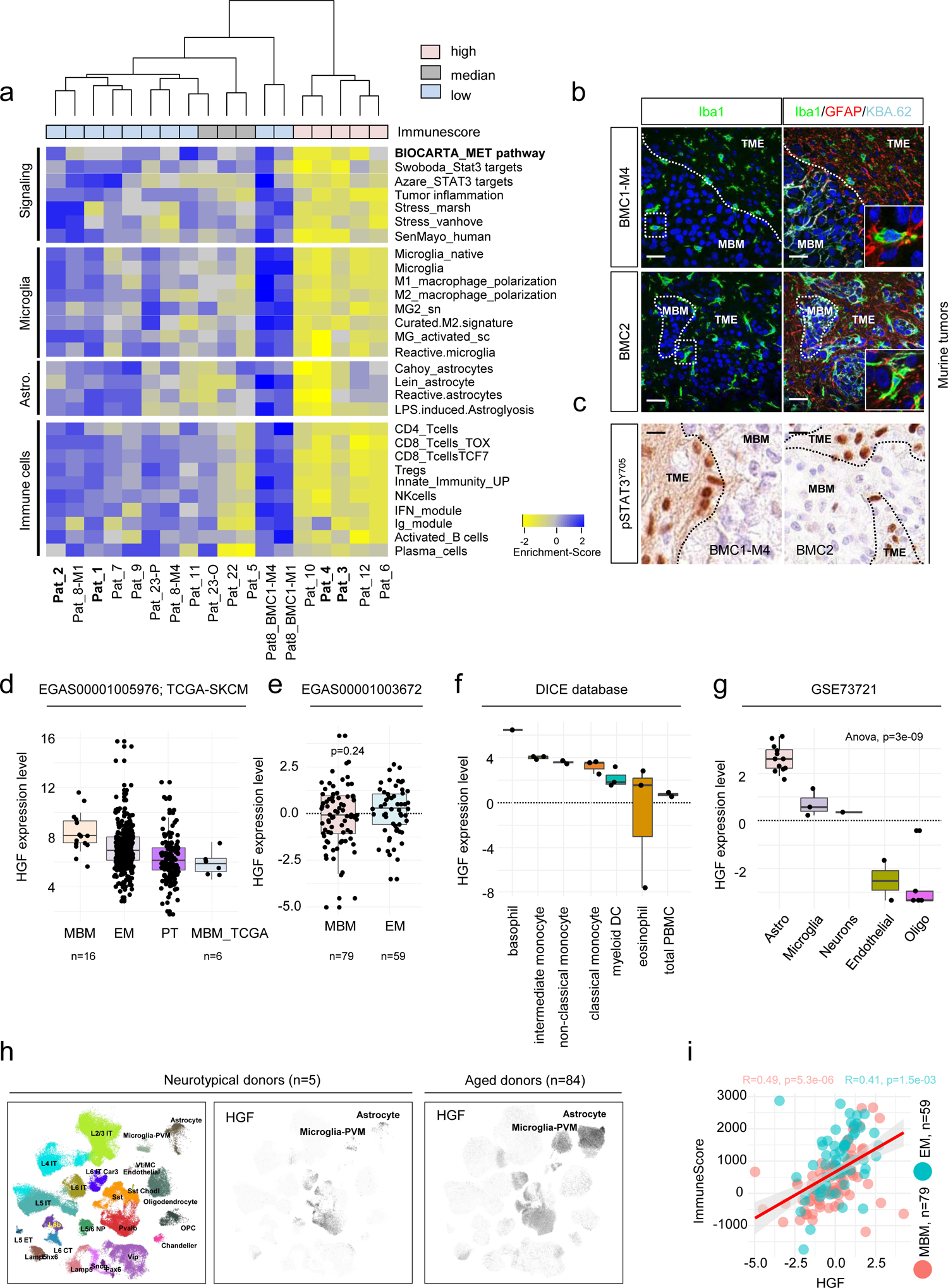
Signature-based deconvolution identified parameter of MBM featuring a favorable disease course and identified a role of MET signaling. a.) Single-sample GSEA (ssGSEA)-based deconvolution of MBM of study EGAS00001005976 using customized gene signatures indicating “Signaling” processes, cellular subsets and stages of microglia and astrocyte and immune cell subsets. ssGSEA demonstrated distinct separation of MBM with high, median or low immune score regarding expression levels of signature genes, BMCs served as controls. ssGSEA uncovered differentially activated pathways and processes such as MET and STAT3 and interferon signaling, senescence (SenMayo), stress response and tumor inflammation in tumors enriched for reactive microglia and astrocytes and innate and acquired immune cells subsets. b.) Confocal microscopy images of orthotopic tumors established by stereotactic injection of BMC1-M4 or BMC2 cells into brains of Crl:CD1-Foxn1^nu^ mice^23^, stained for Iba1 (green, microglia) or Iba1, GFAP (red, astrocytes) and KBA.62 (turquoise, pan-melanoma cell marker). DAPI served as nuclear dye. Markers show distinct areas of tumor (MBM) and microenvironment (TME) and regions of microglia infiltration, 21 days after intracranial injection^23^. MBM-TME boarders are indicated by white, dashed lines. c.) IHC of tumors investigated in (a) for activation and tyrosine phosphorylation (residue Y705) of STAT3. pSTAT3^Y705^ is particularly present in microenvironmental cells (astrocytes). Black, dashed lines indicate MBM-TME boarders. In b, c, bars indicate 50 µm. d-e.) Expression levels of hepatocyte growth factor (HGF) in tumors of studies EGAS00001005976, TCGA-SKCM and EGAS00001003672 demonstrating HGF expression in all tumor subsets. f-g.) Investigation of HGF expression in immune cell subsets (DICE database^35^) and brain cells (study GSE73721) revealed highest levels in basophil granulocytes and monocytes (f) and in astrocytes and microglia (g). h.) UMAP projection of expression profiles from nuclei isolated from 5 neurotypical donors as provided by Seattle Alzheimer’s disease brain cell atlas (https://portal.brain-map.org/explore/seattle-alzheimers-disease), cellular subtypes are color coded (left panel). Log-normalized expression levels of HGF in nuclei isolated from 5 neurotypical donors (center panel). Log-normalized expression levels of HGF in nuclei isolated from 84 aged donors (42 cognitively normal and 42 with dementia), right panel, demonstrating increased number of HGF expressing microglia and astrocytes as triggered by inflammatory processes. i.) Dot plot showing the correlation of HGF expression and immune score of BM (R=0.49, p=5.3e-06) and EM (R=0.41, p=1.5e-03) indicating a potential role of HGF in immune cell-related processes. Box and whisker plots show median (center line), the upper and lower quartiles (the box), and the range of the data (the whiskers), including outliers (d-g). Significance was determined by unpaired, two-sided t-test (e) or one-way ANOVA (g).

HGF or scatter factor (SF) is the only identified ligand of MET, plays a pivotal role during neural development, regulating growth and survival of neurons^15,44^ and likely serves as inducer of reactive microglia by an autocrine loop in response to trauma or neurodegenerative disorders^13^. Therefore, MET expressing, brain colonizing melanoma cells may benefit and take advantage of the HGF-controlled systems naturally occurring in the brain. We observed HGF expression among tumors of different data sets comprising MBM, EM and primary tumors (studies EGAS00001005976; TCGA-SKCM; EGAS00001003672) with no significant difference of HGF levels in tumor subsets (Figures 3d, e). Investigation of immune cell and brain cell data (DICE database^35^ and study GSE73721^45^) revealed high expression of HGF in monocytes and astrocytes (Figures 3f, g), suggesting a potential role of different stroma cell populations for activating HGF/MET signaling in brain-infiltrating tumor cells. Assuming that the degree of microglia infiltration determines signaling processes in MBM cells, we explored expression levels of MET signaling-associated genes in tumors with high and low level of infiltrated TAMs and found levels of HGF, PIK3CG, PTK2B, STAT3 and MAP4K1 significantly correlated with microglia score (Supplementary figure 4c-e) that was defined as average expression level or β-value of microglia markers *APBB1IP, SYK, HCK* and *P2RY12* (Supplementary table 6).

HGF might be released by immune cells as well as homeostatic and reactive microglia or astrocytes. We surveyed the Seattle Alzheimeŕs Disease Brain Atlas which is implemented in the Allen brain atlas database (https://portal.brain-map.org/) and observed that dementia fostered expansion of microglia with increased expression of HGF (Figure 3h, center and right panels). Reactive microglia and immune cell released HGF might hence be responsible for activation of growth factor/survival signaling in adjacent tumor cells. In line with previous studies, we observed a significant correlation of HGF expression with immune score in brain (BM, R=0.49, p=5.3e-06) and extracranial metastases (EM, R=0.41, p=1.5e-03), (Figure 3i).

### Expression and activation of MET receptor classifies a molecular subset of MBM

Understanding the molecular mechanisms that establish cellular dependencies and thus control the development and maintenance of brain metastases is critical for their therapeutic manipulation. Recently, we identified that the expression of Ecad and NGFR sufficiently discriminated molecular subsets of MBM^23^. These subsets likely distinctly interact with microenvironmental cells and respond to therapeutics (Figure 4a). To identify potential druggable targets, we surveyed the pan-MBM, NGFR and Ecad-specific gene sets for cell surface receptors that may serve as crucial key factors that control tumor cell maintenance and expansion and identified 24 receptors that distinguished Ecad^+^ and NGFR^+^ tumors (Figure 4b). Particularly ADIPOR1 (adiponectin receptor 1, p=1.9e-02), SIRPA (signal regulatory protein alpha, p=1.1e-05) and PLXNC1 (plexin C1) showed significantly increased expression in Ecad^+^ MBM and EM but comparable levels among MBM and EM (Figure 4c). In addition, Ecad^+^ MBM featured increased levels of MET receptor in (p=1.4e-04). MET was significantly (p=2.7e-05) higher expressed in MBM than EM (Figure 4d, left and center panels). The MET tyrosine kinase receptor pathway serves as a potent survival and maintenance factor for MBM and might be a promising therapeutic target^46^. MET expression was associated with increased cell cycle progression and proliferation (Figure 4d, right panel) and defined yet another subset of MBM (Figure 4e). Next we assessed whether expressed MET indeed participated in active signaling processes. Phosphorylation of MET at tyrosine residues 1234/1235 (pMET^Y1234/1235^) is critical for kinase activation and initiation of downstream processes and was evident in nearly all MET^high^ MBM investigated, independent of the BRAF mutation status (Figures 4f, g). MET receptor alterations are evident in 9 % of all SKCM melanoma cases, including amplification as observed in 1.13 – 17.2% or 11% of melanoma (TCGA-SKCM, study by Ramani et al.^47^). However, targeted DNA sequencing (TargetSeq) and fluorescence in-situ hybridization (FISH) revealed absence of MET activating mutations and a tendential MET amplification in only one case (Pat 5, Supplementary figure 5a, b). However, all but one tumor (Pat 14) showed high polysomy. As we assume that environmental cells foster activation of MET receptor signaling in a subset of tumor cells, we performed co-IHC for pMET^Y1234/1235^ and Iba1. We observed pMET^Y12^^34/1235^ positive tumor cells in close proximity to Iba1^high^ TAMs (Figure 4h), though MET receptor was not activated in Iba1^high^ microglia that resided in adjacent normal tissue (Supplementary figure 5c, upper panel). However, MET receptor activation was also evident in scattered tumor cells in the absence of adjacent Iba1^high^ TAMs (Supplementary figure 5c, lower panel) suggesting paracrine mechanisms or additional sources of HGF such as immune cells or astrocytes. Considering that HGF levels, like those of other growth factors provided by stromal cells, might depend on spatial factors, we examined the Allan Brain Atlas database and found that HGF is comparably expressed in different brain sections (frontal lobe (FL), parietal lobe (PL), temporal lobe (TL), occipital lobe (OL)) but is lowly abundant in the brainstem (pons) (supplemental Figure 5d). The spatially dependent expression of growth factors in the brain may therefore determine the dependencies of the tumor cells.

**Figure 4:**
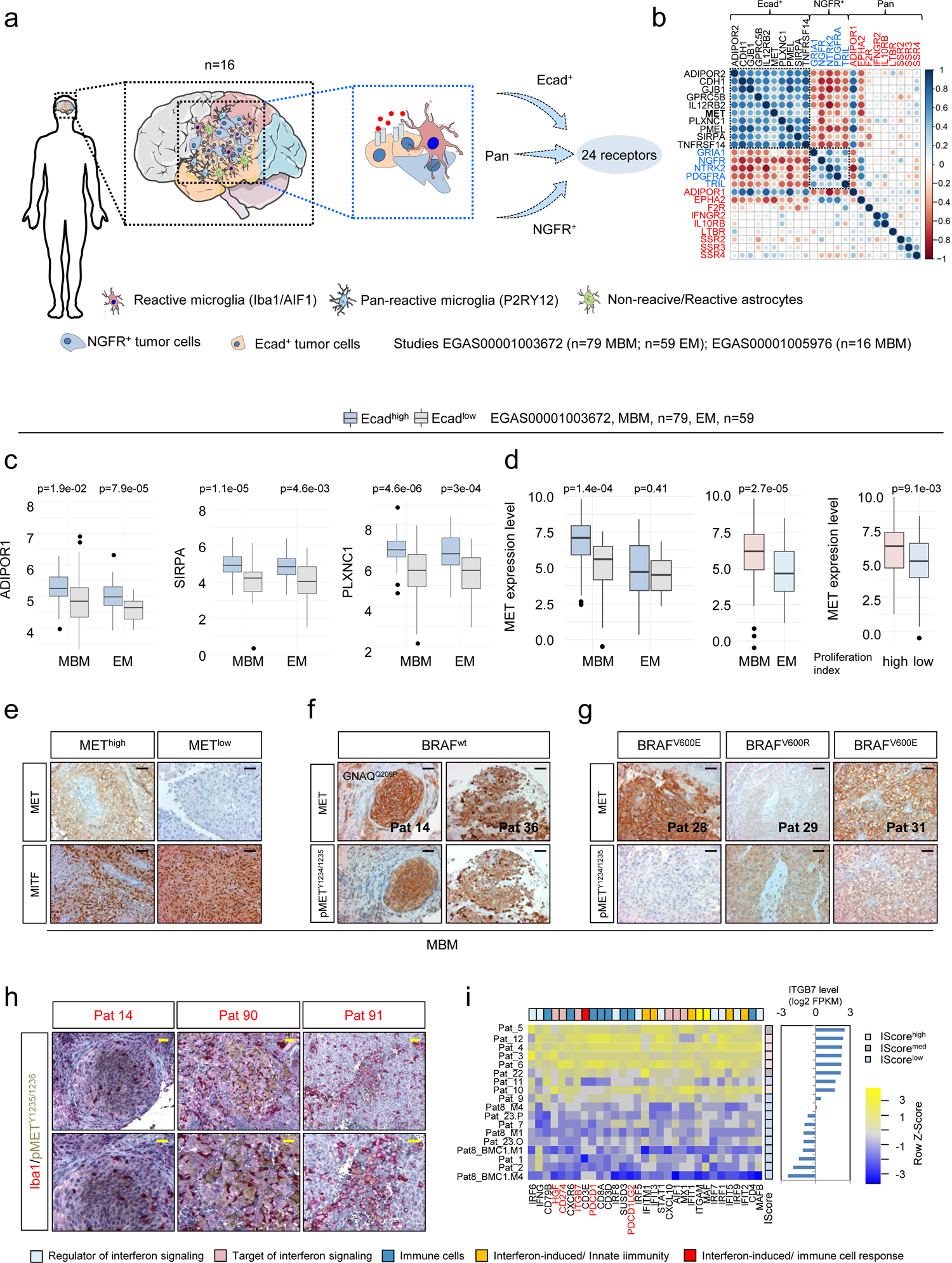
Ecad^+^ MBM are defined by expression of MET receptor. a.) Schematic summary of the initial screen of MBM expression data of our recent study (EGAS00001005976; n=16 MBM) for subset expressed receptors. MBM contain Ecad^+^ and NGFR^+^ subsets and admixed cells such reactive microglia, labeled by expression of Iba1/AIF1 and or P2RY12. The initial survey yielded 24 receptors that potentially establish cell survival/growth of MBM. b.) Correlation map (Spearman, p<0.05) showing the relationship of identified receptors expressed in MBM of our previous study, emphasizing the distinct pattern of Ecad^+^ and NGFR^+^ molecular subsets. The value of the correlation coefficient is color coded. c.) Box plots depicting the levels of most relevant receptors that significantly separated Ecad^high^ and Ecad^low^ subsets of MBM and extracerebral metastases (EM) of study EGAS00001003672, providing MBM = 79, EM = 59 (ADIPOR1, Adiponectin Receptor 1, p=0.019/7.9e-05; SIRPA, Signal Regulatory Protein Alpha, p=1.1e-05/0.0046; PLXNC1, Plexin C1, p=4.6e-06/3e-04). d.) Box plots depicting the levels of MET in Ecad^high^ and Ecad^low^ subsets of MBM and EM (left panel, p=1.4e-04/p=0.41) or in all subtypes of MBM and EM (p=2.7e-05) in high and low proliferating tumor cell subsets (right panel, p=9.1e-03). e.) IHC of selected MBM for MET and MITF validated the two subsets. f-g.) Expression and activation status of MET in BRAF wildtype (wt; Pts 14, 39) and BRAF^V600E/R^ mutated MBM (Pts 28, 29, 31). Phosphorylation of MET at residues Y1234/1235 is critical for kinase activation. h.) IHC of indicated tumors for co-localization of pMET^Y12^^34/1235^ (brown) and Iba1 (red) demonstrating potential activation of MET receptor signaling tumor cells by stromal cell-secreted HGF. i.) Heat map representing expression levels of regulators and targets of interferon signaling and immune related genes showing clustering according to the level of ITGB7 expression. Box and whisker plots show median (center line), the upper and lower quartiles (the box), and the range of the data (the whiskers), including outliers (c, d). Significance was determined by unpaired, two-sided t-test (c, d).

### Interferon signaling determines response of MBM to immune checkpoint inhibitor therapy

Interferon-gamma signaling has been identified as an important mechanism for upregulation of PD-L1 on melanoma cells and escape from immune recognition. On the other hand, recent studies uncovered that high interferon-gamma-related gene expression signature scores (IFN-γ score) were associated with low risk of melanoma relapse from neoadjuvant ipilimumab plus nivolumab therapy^48,49^.

In our recent study, we observed significant enrichment of interferon and inflammatory response (“Hallmark”, MsigDB^50^) signatures in MBM with high level of tumor infiltrating lymphocytes (TIL^high^)^23^ that have been attributed with favored survival in a pre-clinical melanoma model^49^. We found overlapping expression of *ITGB7*, *SUSD3* and *HGF* and Hallmark interferon-response genes, separating MBM of our cohort and MBM of study EGAS00001003672 (Figure 4i, Supplementary figure 6a). Expression of *ITGB7* significantly correlated with levels of interferon regulatory factor 1 (IRF1) and IRF8 in MBM (BM) and extracranial metastases (EM) of study EGAS00001003672 (Supplementary figure 6b-d). Moreover, we observed high correlation of levels of *HGF*, *IRF1* and *IRF8* in MBM (Supplementary figure 6e). As microglia serve as a source of soluble receptor ligands such as Hgf, we next surveyed data of interferon-gamma treated (1 U/mL IFNγ, 24h) murine microglia cells (BV2, GSE132739). Indeed, we found significant upregulation of *Itgb7* (p = 2.9e-03) and *Hgf* (p=4.4e-02) but downregulation of *Susd3* (p=4.0e-02) in BV2 cells (Supplementary figure 6f). For control, we investigated levels of known interferon-responsive genes that were significantly increased 24h after interferon treatment, *Mx1* (p=1.2e-02), PD-L1/*Cd274* (p=3.6e-02), *Irf1* (p=3.1e-02) *Cxcl9* (p=4.0e-03) and *Aif1* (p=3.9e-04) (Supplementary figure 6g). In order to classify MBM of our study into anti-PD-L1 responsible and non-responsible and for linking *ITGB7*, *SUSD3* and *HGF* with therapy response, we performed ssGSEA and applied interferon responsive and additional immune response gene signatures (of study GSE186344^51^). Our survey validated that *ITGB7*, *SUSD3* and *HGF* were highly expressed in MBM that featured enrichment of interferon responsive genes/signatures (Pts. 3-6, 12; Supplementary figure 6h). Hence, we suggest that *ITGB7*, *SUSD3* and *HGF* like *PD-L1* are among the interferon-regulated genes triggered by immune cell-released interferon-gamma and may be involved in immune response mechanisms of MBM.

### The targeting of MET receptor serves as a promising strategy to control MBM growth

Although a subset of MBM exhibit immune cell subset enrichment and interferon response signatures and respond to ICi therapy, MET-expressing brain metastatic melanoma cells may benefit from HGF released by stromal cells to drive progression. Hence, activation of MET signaling may depend on the degree of tumor-stroma interaction, possibly counteracting the beneficial impact of immune checkpoint inhibition (ICi). Resistance-mediating processes include the phosphorylation of ribosomal protein S6 (pS6), which is downstream of MET and mTOR signaling^52^ and was observed in progressive BRAFi-resistant melanomas^53,54^.

We assessed pS6 phosphorylation of serine residues 235/236 and found co-occurrence of activated MET receptor and of pS6^2^^35/236^ in MITF positive tumors (Figure 5a and Supplementary figures 7a, b). Moreover, pS6 phosphorylation was evident in a BRAF^wt^ (T2002) and mutated (V600E, BMC53) cell lines probably suggesting a general activation of pS6 signaling irrespective of the presence of mutated BRAF (Figure 5b). As MET signaling might serve as mediator of a resistance-mediating program, we assessed the efficacies of the ATP-competitive inhibitor PHA-665752 and the non-ATP-competitive, clinical phase I/II MET receptor inhibitor (METi) tivantinib (ARQ197) in BMCs that showed variable levels of MET expression (Figure 5c and Supplementary figure 7c). ARQ197 failed to improve the outcome and overall survival of patients with hepatocellular carcinoma^55^ but may potentially be effective in melanoma patients. The initial testing revealed a general response of BMCs (BMC1-M1, BMC53), T2002 cells and conventional cell lines (A375, A2058, MeWo) to both inhibitors irrespective of the BRAF mutation status (Figures 5d, e). As we observed a mutually exclusive rather than co-expression of MET receptor and NGFR (nerve growth factor receptor), we tested whether the manipulation of NGFR levels might affect the response to PHA-665752 (PHA). We observed that overexpression of NGFR in A375 cells (A375^NGFR^) sensitized to METi compared to RFP expressing control (A375^RFP^) or MeWo cells (Figure 5f). Next, we asked whether METi targeting may serve as alternative therapeutic strategy for BRAFi resistant (BMC4) or cells with non-BRAF^V600^ mutations (BMC2) showing only moderate or no response to dabrafenib (Figure 5g) as indicated by IC_50_ values (BMC4, IC_50_ = 226.4 nM and BMC2, IC_50_ = 3029.5 nM). The broad range (1nM – 10µM) testing of tivantinib in BMCs, T2002 and conventional melanoma cells and PHA in BMCs revealed that all cell lines responded to both METi, irrespective of the mutation status. However, we observed that the non-(brain) metastatic cell lines A375, A2058, and T2002 were more sensitive to treatment with tivantinib than BMCs (Figures h-j, Supplementary figure 7d). The median IC_50_ value of BMCs was ∼600 nM (range: 406.5 – 800.1). Cell lines lacking BRAF and NRAS mutations (MeWo, T2002) showed highest responses to tivantinib (Figure 5k).

**Figure 5:**
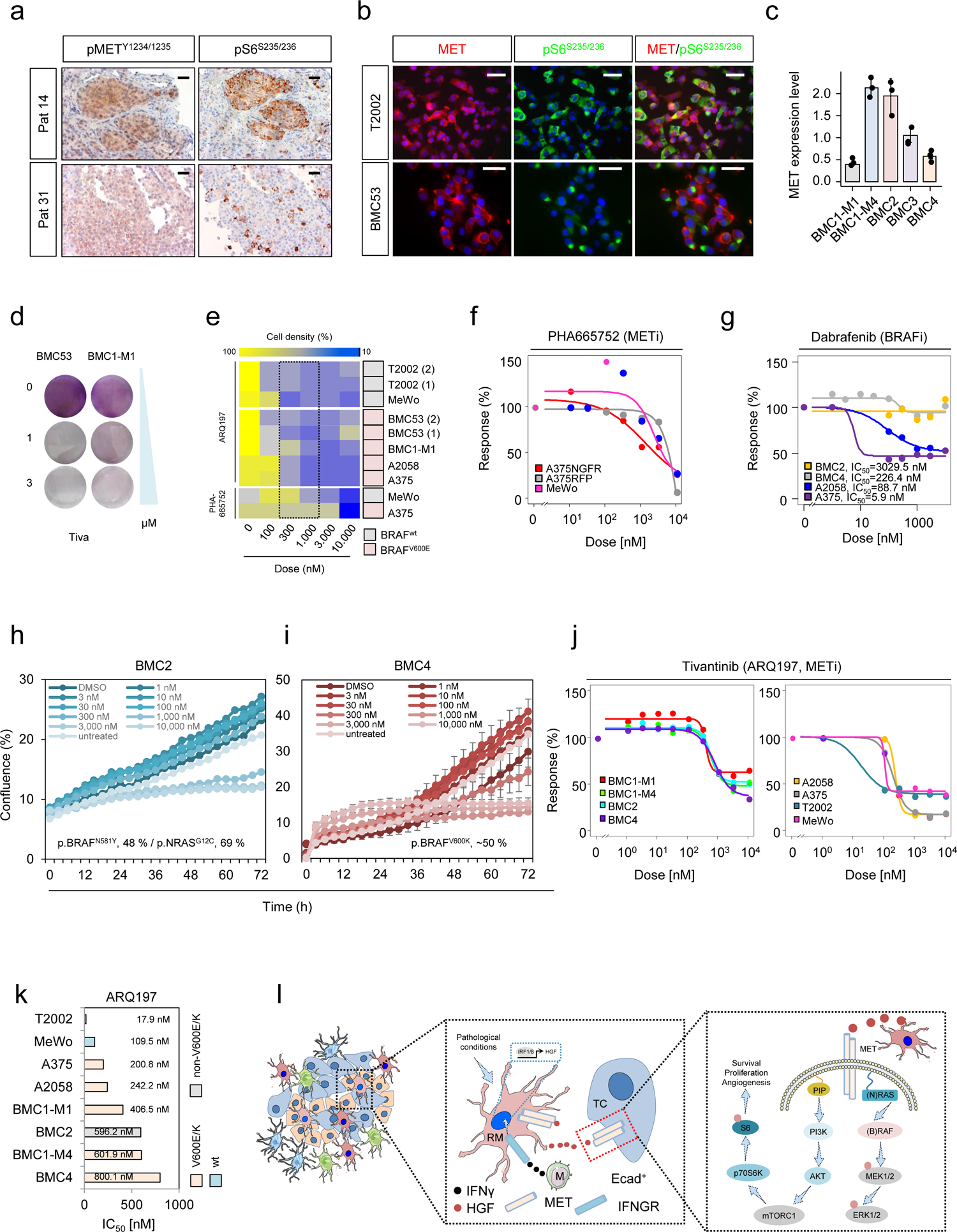
Inhibitors of MET receptor decrease growth of brain metastatic and conventional melanoma cell lines. a.) Comparative IHC of selected MBM for levels of phosphorylated and activated MET receptor (pMET^Y12^^34/1235^) and ribosomal protein S6 (pS6^2^^35/236^) of consecutive sections suggesting MET-associated activation of mTOR signaling. b.) Immunofluorescence microscopy of lymph node-metastatic (T2002) and brain metastatic (BMC53) patient-derived melanoma cell lines for co-occurrence of MET (red) and pS6^2^^35/236^ (green). DAPI served as nuclear dye. c.) qPCR analysis of BMCs for expression of MET receptor, bars indicate median levels ±SD of three biological replicates. d.) Gross initial ARQ197 sensitivity test of BMC53 and BMC1-M1 cells showing high and low levels of MET expression. Cell density was determined by crystal violet staining. e.) Broad range determination of sensitivity of BMCs, T2002 and conventional melanoma cell lines (A375, A2058, MeWo) to METi PHA-665752 and ARQ197. Cell density and BRAF mutation status are indicated. Dotted line depicts the estimated range of IC_50_. f.) PHA-665752 dose-response fit curve-based calculation of IC_50_ values of A375 cells with overexpression of NGFR or RFP control cells and MeWo cells. g.) Dabrafenib dose-response fit curve-based calculation of IC_50_ values of BMCs exhibiting different BRAF mutations (BMC2^p.N581Y^, BMC4^p.V600K^) and A375^p.V600E^, A2058^p.V600E^ cells. h-i.) Live cell imaging-based tracking of confluence (%) of BMC2 and BMC4 cells in dependence of increasing doses of ARQ197. Shown are median values±SD of eight technical replicates. A representative out of two experiments is shown. j.) ARQ197 dose-response fit curves of BMCs, T2002 and conventional cell lines. Calculated IC_50_ values are indicated, suggesting response of dabrafenib resistant cell lines to METi. k.) Bar diagram summarizing IC_50_ values (nM) indicating the response of indicated cell lines to ARQ197. The BRAF status is color coded. l.) Working model suggesting the activation of MET receptor signaling in adjacent tumor cells and in (reactive) microglia (RM) by microglia released HGF. IRF-mediated HGF expression in turn is triggered by immune cell (monocytes/macrophages, M) released interferon-gamma. HGF binding to tumor cell (TC) expressed MET receptor directs downstream activation of the RAS/RAF/MEK/ERK and the PI3K/AKT/mTOR/p70S6K branch. The latter is leading to phosphorylation and activation of the ribosomal protein S6. Box and whisker plots show median (center line), the upper and lower quartiles (the box), and the range of the data (the whiskers), including outliers (c).

In summary, brain metastatic as conventional melanoma cell lines responded to METi, suggesting that targeting of MET signaling might be a promising tool for the treatment of non-BRAF^V600^ and BRAF^V600^ mutated MBM that acquired resistance to BRAFi or for combinatorial of METi and ICi in NRAS mutated tumors.

## Discussion

The spatiotemporal development of primary and secondary brain tumors is strongly determined by the crosstalk of tumor and brain micronenvironmental cells, particularly macrophages, astrocytes and microglia^56^ and the consequential activation of inflammatory processes^57^. Although the neuro-inflammatory processes that are activated alongside development and progression of primary brain tumors such as glioblastoma have been intensively studied, the mechanisms that accompany emergence of brain metastases on the other side are not well investigated.

Here, we used combined transcriptome and methylome profiling to unravel the molecular features of MBM of different progression stages showing high and low level of tumor-associated macrophages/microglia (TAMs) infiltration, irrespective of the phenotype (Ecad, NGFR). Generally, TAMs foster development and progression of primary brain tumors^58,59^, however their functional role in MBM may be different. We observed that MBM containing a high proportion of TAMs were associated with a high immune score and infiltration of CD3^+^ T cells. The profiling of Iba1/AIF1^high^ tumors revealed a cluster of genes, among them *ITGB7*, *APBB1IP* as *SUSD3* and *PD-L2*, that were widely expressed among immune cell subtypes and previously associated with increased immune T cell infiltration^23,27,28,33^ and favored outcome. Previous mouse studies demonstrated a pivotal role of ITGB7 for intestinal T cell recruitment and correlated low levels of Itgb7 with colorectal cancer progression and maintenance of intestinal stem cells via Ecad-mediated interaction^27,33^. However, we observed strong protein expression of ITGB7 in immune cells adjacent to tumor cells of Ecad^+^ and NGFR^+^ tumors, suggesting a broader function of ITGB7 in different subtypes of metastases and cancers. Enhanced expression of ITGB7 might be a prerequisite for immune cell invasion. Therefore, epigenetic marks that correlate with the expression of ITGB7 and other genes mentioned above may be of prognostic importance, and the expression of these markers could determine the pathways of intracranial progression.

As previously described for the Ecad^+^ and NGFR^+^ subtypes of MBM, whether tumors are enriched or depleted in TAMs and immune cell subsets is critical and may determine response to therapeutic intervention. The subsequent ssGSEA-based characterization of MBM of studies performed by us and others revealed molecular programs fostering or accompanying the TAM^+^/TIL^+^ tumor subtype. TAM^+^/TIL^+^ tumors featured activation of MET and STAT3 signaling, increased stress response, tumor inflammation, senescence and activation of microglia and astrocytes but also activated interferon signaling. STAT3 activation in tumor-adjacent astrocytes in response to brain damage or tumor cells is well-investigated process^60,61^ and was rapidly induced in response to brain infiltrating BMCs. Hence, enrichment of STAT3 signature genes was likely attributed to tumor-adjacent astrocytes and infiltrated immune cells^62^.

The HGF/MET receptor signaling plays a pivotal role during brain development and neuro-regeneration, homeostasis of microglia and neurons^14,44^ but is also involved in microglia activation in response to trauma^13,44,63^. Brain infiltrating melanoma cells hence may engage the HGF/MET signaling of brain cells and utilize for regulation of survival and proliferation. We observed expression of MET receptor in the subset of E-cadherin (Ecad) expressing tumors^23^, suggesting that Ecad^+^ but not NGFR^+^ cells may depend on HGF/MET signaling. As we observed that HGF is expressed by immune cell subsets and homeostatic or reactive astrocytes and microglia, we investigated the level of activated/phosphorylated MET receptor in TAM-adjacent tumor cells. We found that tumor cells but not Iba1^high^ TAMs that resided in tumor cell-free adjacent stroma showed activation of MET, however MET was also activated in the absence of adjacent microglia in some tumor cells, suggesting a paracrine effect of HGF. In line with our previous study^23^, we observed enrichment of interferon-response signatures in the subset of TIL^high^/immune score (IS)^high^ tumors. We observed enrichment of interferon-response genes in MBM with high levels of ITGB7 expression and observed significant response of Itgb7 and Hgf among known interferon-inducible genes such as Cd274^64^ and Mx1^65^ in interferon-gamma treated BV2 murine microglia cells (unpublished study GSE132739). Hence, T cell-provided interferon-gamma might not only induce expression of Cd274/PD-L1 but may also activate expression of HGF and ITGB7. Therefore, autocrine MET receptor signaling might be triggered in response to immune cell released interferon-gamma and/or paracrine activation of MET signaling may occur via (INFG-activated) reactive glia-released HGF.

Our study bridges the gap between the immune cell phenotype of MBM and the activation of potentially therapeutic counteracting signaling pathways. The infiltration of TAMs and immune cells thus represents a double-sided sword and on the one hand is associated with an effective response to immune checkpoint inhibitors, but on the other hand can support the growth of MET expressing tumor cells via secreted factors such as HGF.

Therefore, we finally assessed the potential role of small molecule inhibitors of MET receptor (METi) for targeting of MBM that lack druggable BRAF^V600^ mutations or developed refractory disease. To this end, we took advantage of well-characterized BMCs serving as *in vitro* model systems. We observed that the ATP-competitive inhibitor PHA-665752 and the non-ATP-competitive, clinical phase II inhibitor ARQ197 (tivantinib) elicited response in BMCs and conventional melanoma cell lines irrespective of the BRAF/NRAS mutation status. However, although being effective at doses of 100 – 200 nM in MeWo and A375 cells, ARQ197 showed a median IC_50_ value of ∼1 µM in BMCs, suggesting a general difference among brain metastatic and long-term maintained conventional cell lines established from either non metastatic (A375) or locally metastatic (MeWo) cells.

In summary, we have shown that MET receptor signaling is active in a subset of MBM, conferring a survival/growth benefit independent of BRAF/NRAS mutation status. MET activation may occur in response to HGF released by TAM/immune cells and could counteract therapeutic interventions. Furthermore, we suggest interferon-induced expression of HGF in tumor cells triggered by interferon-gamma provided by stromal cells mediates autocrine activation of MET-signaling tumor cells (Figure 6). In addition, we demonstrated that methylome profiling of MBM has high potential to identify gene regulatory sites that may predict favorable progression of intracranial disease. In the present study, we identified epigenetic regulatory sites in a group of genes comprising *ITGB7*, *APBB1IP*, *SUSD3* and PD-L2 (*PDCD1LG2*).

**Figure 6:**
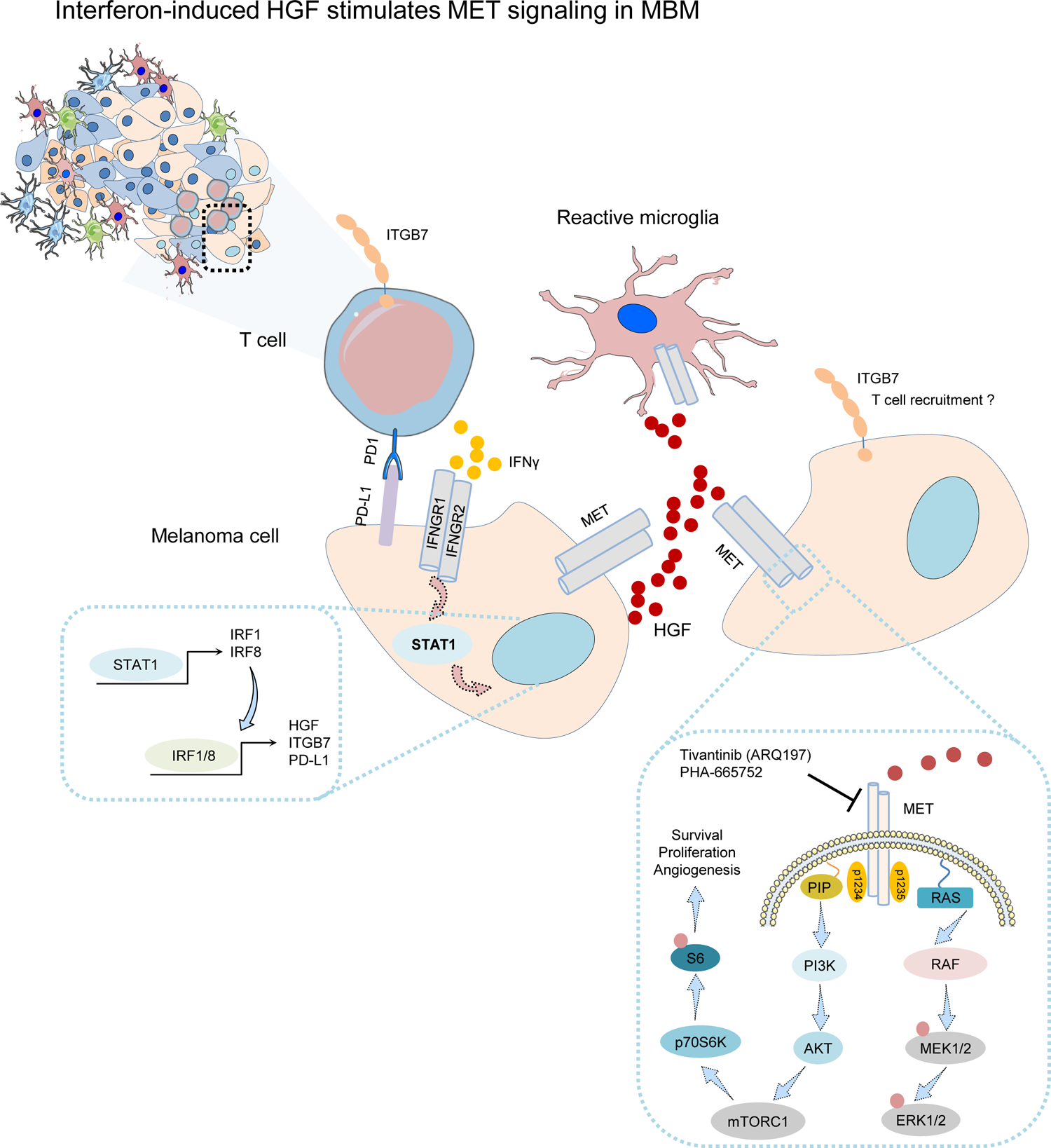
HGF/MET receptor signaling might be activated in tumor cells at immune cell/TAM dense areas. Schematic representation of our working model suggesting the interaction of tumor cells with stromal cells, particularly microglia and immune cells, consequentially leading to activation of MET signaling in tumor cells via stromal cell-released HGF. Expression of HGF in turn and ITGB7 and PD-L1 is likely triggered by T cell provided interferon-gamma. Increased levels of ITGB7 may foster recruitment of immune cells.

### Limitations of the study

The present study is not without limitations. Although we suggested that HGF/MET signaling is activated in tumor cells in close proximity to infiltrated microglia, we have not provided evidence that growth of established brain tumors and phosphorylation of MET decreases in response to METi. Moreover, whether METi are capable of passing the blood-brain barrier and not affect normal homeostatic processes e.g. those crucial for neuron survival needs to be investigated.

## Methods

### Patient cohorts

All procedures performed in this study were in accordance with the ethical standards of the respective institutional research committees and with the 1964 Helsinki declaration and its later amendments or comparable ethical standards. All patients gave written informed consent for the collection and scientific use of tumor material which was collected at the Biobank of the Charité – Comprehensive Cancer Center (CCCC). The study was approved by the Ethics Committee of the Charité (EA1/152/10; EA1/107/17; EA4/028/18 and EA1/107/17 and EA1/075/19) and Universitätsmedizin Greifswald (BB 001/23).

### Cultivation of MBM-derived and conventional melanoma cell lines

All cell lines were cultured as previously reported^23^. Briefly, all brain metastases-derived cell lines (BMCs) and conventional melanoma cell lines were kept at 37°C/ 5% CO_2_ and 95% humidity in cell culture medium (DMEM, 4.5 g/L glucose, stabilized glutamine/GlutaMax, pyruvate, Gibco/ThermoFisher) supplemented with 10% fetal bovine (FBS, Gibco) serum and 1% penicillin/streptomycin (P/S) (Gibco/ThermoFisher) and routinely passaged. BMCs were established from intraoperative tumors as previously reported^23^.

### Live cell imaging-based drug sensitivity assays

Drug treatments were performed 24 h after seeding of 2,500-5,000 cells/96-well in 100 µl medium. The response of BMCs and conventional melanoma cell lines to dabrafenib, PHA-665752 or ARQ197 (all purchased from Selleckchem) in a range of 1nM-10µM of eight technical replicates was determined by live cell imaging. Images were taken every three hours using a 10x objective and the general label-free mode, two pictures of eight technical replicates per condition were taken. Drug response was assessed by changes in the cellular density over time. The cell density was determined by a confluence mask tool as part of the IncucyteS3 software. IC50 values were calculated by curve-fitting (https://search.r-project.org/CRAN/refmans/REAT/html/curvefit.html) based on confluence measurements at day 3.

### In vivo experiments

All animal experiments were performed in accordance with the German Animal Protection Law under the permission number G0130/20 obtained via the Berlin Ministry of Health and Social Affairs (LaGeSo). ARRIVE 2.0 Guidelines were strictly followed and performed as previously reported^23^. Briefly, 2.5×10^4^ BMC1-M4 and BMC2 cells were stereotactically inoculated into brains of female Crl:CD1-Foxn1^nu^ nude mice (8-9 weeks of age, 24-26g, Charles River Laboratories) were with using a 1µl Hamilton syringe and a stereotactic frame as described previously^66^. Tumor growth was tracked by MRI and animals were sacrificed by perfusion with 4% PFA in deep anesthesia after tumors reached a volume of 20 mm³. Following, whole brains were removed, dehydrated, paraffin embedded and sections of 2 µm were used for downstream analyses.

### RNA isolation and sequencing

Isolation of total RNA from snap frozen tumors and RNA sequencing was performed as previously reported^23^. Briefly, 100 ng of total RNA was used for library preparation with TruSeq Stranded total RNA Sample Preparation-Kit and Ribo-Zero Gold Kit (Illumina). Paired-end (2×100 bp) sequencing of RNA libraries with integrity numbers (RIN) ≥7 was performed on NovaSeq6000 platform at Cegat GmbH, Tuebingen (Germany). Following demultiplexing of sequenced reads and adapter trimming^67^, FASTQ files were obtained. Raw counts of protein-coding genes were normalized using the DESeq2 (https://bioconductor.org/packages/release/bioc/html/DESeq2.html) package^68^. Differential expression of genes between groups was determined after fitting models of negative binomial distributions to the raw counts. Raw p-values were FDR (false discovery rate)-adjusted for multiple testing and a value below 0.05 for the adjusted p-values were used to determine significant differentially expressed genes.

### Gene-set enrichment GSEA/Single-sample GSEA/Scores

GSEA was performed using the most current BROAD javaGSEA standalone version (http://www.broadinstitute.org/gsea/downloads.jsp) and gene signatures of the molecular signature database MsigDB^50,69^, v7.4. In addition, we performed GSVA/ssGSEA using R packages GSVA^70^, GSRI, GSVAdata and org.Hs.eg.db and a customized collection of gene signatures including the signatures provided by Biermann et al.^71^ and own signatures as defined by selected Ecad^23^, NGFR^23^, microglia or TME core genes (this study). All gene signatures are shown in Supplementary table 5. Microglia scores were defined as the mean β-value of probes cg24400465 (APBB1IP), cg05128364 (SYK), cg21704050 (P2RY12) and cg03498995 (HCK) or expression levels (log2 FPKM) of these markers. The proliferation index in Figure 4d was defined as mean expression level of the cell cycle regulators PCNA, MKI67, CCNB1 and CCNB2.

### Fluorescence in situ hybridization (FISH)

FISH analysis was performed on 4 µm sections of FFPE blocks. Slides were deparaffinized, dehydrated and incubated in pre-treatment solution (Dako, Denmark) for 10 min at 95–99°C. Samples were treated with pepsin solution for 6 min at 37°C. For hybridization, a Vysis MET SpectrumRed/ Vysis CEP 7 (D7Z1) SpectrumGreen Probe (Abbott, Chicago, USA) was used. Incubation took place overnight at 37°C, followed by counterstaining with 4,6-diamidino-2-phenylindole (DAPI). For each case, signals were counted in 50 non-overlapping tumor cells using a fluorescence microscope (BX63 Automated Fluorescence Microscope, Olympus Corporation, Tokyo, Japan). Computer-based documentation and image analysis was performed with the SoloWeb imaging system (BioView Ltd, Israel). MET high-level amplification (MET FISH+) was defined as (a) MET/CEN7 ratio ≥2.0, (b) average MET copy number/cell ≥ 6 or (c) ≥10% of tumor cells with ≥15 MET copies/cell as described in Schildhaus et al^72^.

### Quantitative real-time RT-PCR

RNA isolation from frozen cell pellets was performed with the RNeasy Mini Kit (Qiagen, Germany) and, following the manufacturers protocol as previously reported^23^.qRT-PCR was carried out on a Step one plus PCR cycler (Applied Biosystems, Germany) for 30–40 cycles. Primers were designed for 55–60°C annealing temperatures. Relative expression levels were calculated with the ΔΔCT method^73^, normalized to β-actin. Primer sequences are shown in Supplementary table 7.

### Immunohistochemistry (IHC)/Immunofluorescence (IF)

Automated immunohistochemical staining was performed on formalin-fixed, paraffin-embedded (FFPE) tissue sections using the BenchMark Ultra (Ventana) autostainer. The following primary antibodies were used: CD3 (anti-CD3ε, Agilent, catalog number: #A045201-2, rabbit, dilution: 1:100), pMET (phospho-MET, Tyr1234/1235, Cell signaling, catalog number: #3077, rabbit, dilution: 1:100), pS6 (phospho-S6 ribosomal protein Ser235/236, Cell signaling, catalog number: #2211, rabbit, dilution: 1:100), IBA1 (IBA1/AIF-1, ionized calcium-binding adaptor molecule 1, Cell signaling, catalog number: #17198, rabbit, dilution: 1:100), ITGB7 (Integrin beta 7, Thermo Fisher, catalog number: #BS-1051R, rabbit, dilution: 1:100) and pSTAT3 (phospho-STAT3, Tyr705, Cell signaling, catalog number: #9145, rabbit, dilution: 1:100) and MITF (clones C5L+LD5, Zytomed, catalog number: Z2161MP, mouse, dilution: 1:100). Primary antibodies were applied and developed using the iVIEW DAB Detection Kit (Ventana Medical Systems) or the ultraView Universal Alkaline Phosphatase Red Detection Kit (Ventana Medical Systems). All slides were counterstained with hematoxylin for 8 minutes. IF of mouse brain sections was performed with IBA1 (IBA1/AIF-1, ionized calcium-binding adaptor molecule 1, Cell signaling, catalog number: #17198, rabbit, dilution: 1:100), KBA.62, NovusBiologicals, catalog number: NBP2-45285, mAb mouse, 1:100; GFAP-AlexaFluor594, BioLegend, catalog number: 644708, mAb mouse.

## Supporting information

Supplementary figure 1: Iba1/AIF1 expression separates MBM

Supplementary figure 2: The indicators of favorable disease course ITGB7, SUSD3 and APBB1IP are broadly expressed among immune cell subsets

Supplementary figure 3: Methylome profiling uncovered epigenetic regulatory sites in the ITGB7 gene

Supplementary figure 4: Single sample GSEA revealed classification of immune molecular subtypes of MBM

Supplementary figure 5: MET-FISH analysis revealed absence of MET receptor amplifications in MBM

Supplementary figure 6: Expression of interferon-related genes is enriched in MBM of ITGB7high/IScorehigh phenotype

Supplementary figure 7: The mTOR/pS6 signaling is activated in MBM

## Data availability

Whole transcriptome and methylome data were deposited in the European Genome-Phenome Archive (EGA), under accession numbers EGAS00001005975, EGAS00001005976 (https://ega-archive.org/studies/). The data are available under controlled access. Supplementary tables of our recent study have been deposited at Zenodo (https://doi.org/10.5281/zenodo.10006881). Supplementary tables of our previous study containing a full list of patient’s characteristics (Supplementary table 1) have been deposited at Zenodo (https://zenodo.org/record/7013097 and https://doi.org/10.5281/zenodo.7249214).

## Author contributions

TR performed data analyses, prepared figures and wrote the manuscript; ES collected tumors, established BMCs and performed experiments; KP performed histological analyses; MEW, SN and HWSS performed craniotomy and provided MBM; AV performed experiments/ drug response assays; HR provided expertise on microglia markers; AL performed MET-FISH analyses; SKR provided funding; KJ performed bioinformatics data analysis and expertise on data analysis; JR wrote the manuscript, provided resources and funding. All authors have read and agreed to the published version of the manuscript.

## Acknowledgments

We gratefully thank Cathrin Müller for excellent technical assistance.

## Supplementary figure legends

**Supplementary figure 1:** Iba1/AIF1 expression separates MBM a.) Immunofluorescence (IF) for Iba1 (red) and GFAP (labeling of reactive and normal astrocytes) of a NRAS^Q61R^ mutated MBM (Pat 15) that progressed on treatment with immune checkpoint inhibitors (ICi; ipilimumab) showed strong infiltration of tumor-associated microglia/macrophages (TAMs) and the presence of aggregates of microglia and reactive astrocytes. MBM (without adjacent stromal cells) of Pat 3 showed high infiltration of TAMs. DAPI served as nuclear dye. b.) Box plots representing the levels (FPKM, log2) of Iba1/AIF1 in MBM and BMC of study EGAS00001005976 and MBM and EM of study EGAS00001003672. c.) Per sample representation of expression levels (FPKM, log2) of Iba1/AIF1 in MBM, brain metastases-derived cell lines (BMCs) and brain controls (BC) of studies mentioned in (b). d-f.) Survival analyses of MBM patients (n=67) of study EGAS00001003672 or of TCGA-SKCM study (n=459), featuring high or low level of APBB1IP (d, e) or PD-L2 (PDCD1LG2) expression. Analysis revealed a significant (logrank p=5.5e-07) favorable disease course (HR=0.50, Cox-regression analysis) of *APBB1IP*^high^ melanoma (e) and favorable outcome associated with high levels of PD-L2 expression in MBM (f). Survival of MBM patients was not significantly affected by *APBB1IP* levels (d). g.-m.) Correlation and cell type-specificity of microglia markers *SYK*, *HCK*, *P2RY12* and *AIF1*. n.) Dot plot shows significant (R=0.85, p=1.3e-04) correlation of *ITGB7* expression of MBM (n=16, study EGAS00001005976) and immune score. Box and whisker plots show median (center line), the upper and lower quartiles (the box), and the range of the data (the whiskers), including outliers (b).

**Supplementary figure 2:** The indicators of favorable disease course ITGB7, SUSD3 and APBB1IP are broadly expressed among immune cell subsets. a.) Box plots showing expression levels of ITGB7 among T cell subsets (CD4, T helper cells), CD8 (cytotoxic T cells), NK cells (natural killer cells), cDC (conventional dendritic cells), pDC (plasmacytoid dendritic cells) lymphoid follicle-residing B cells (FollicularB cell subsets), innate lymphoid cells, type 3 (ILC3), mast cells, macrophages (Macro), monocyte subsets (Mono), myofibrils (non-immune related cells), plasma cells (PlasmaB) and tumor-associated macrophages (TAMs) as provided by study GSE146771. b.) IHC of MBM of indicated patients for ITGB7. ITGB7 expression is evident in lymphocyte-enriched areas. c.-d.) Boxplot showing expression levels of SUSD3 in immune cell subsets of (a), and shows levels obtained from DICE (Database of Immune Cell Expression, Expression quantitative trait loci (eQTLs) and Epigenomics), suggesting broad but variable expression among immune cell subsets. e.) Comparative illustration of levels of ITGB7 and SUSD3 as requested from DICE. f.) Box plot showing expression levels of APBB1IP among immune cell subsets of the aforementioned GEO study, suggesting a broad expression among immune cell types. Box and whisker plots show median (center line), the upper and lower quartiles (the box), and the range of the data (the whiskers), including outliers (a, c, d, f).

**Supplementary figure 3:** Methylome profiling uncovered epigenetic regulatory sites in the ITGB7 gene. a.) Schematic representation of the ITGB7 gene, showing exons, intronic regions and sites of epigenetic marks as depicted by indicated probes. ITGB7 expression levels are associated with methylation at sites covered by probes cg26689077 and cg01033299 located within a proximal enhancer-like signature and intronic region close to 5’-UTR. Additional regions as covered by probes cg18320160 and cg11510999 are associated with the BRAF mutation status of tumors. b.-c.) Dot plots showing no significant correlation of methylation status (indicated by β-values) at sites covered by probes cg18320160 and cg11510999 and immune score. d.-e.) Box pots indicating a significant association of β-values, determined by aforementioned probes (cg11510999, p = 3e-06; cg18320160, p= 0.003) and BRAF status (BRAF^V600^ vs. wt/NRAS^Q^^61^) of MBM. Box and whisker plots show median (center line), the upper and lower quartiles (the box), and the range of the data (the whiskers), including outliers (d, e).

**Supplementary figure 4:** Single sample GSEA revealed classification of immune molecular subtypes of MBM. a.) Single-sample GSEA (ssGSEA)-based deconvolution of MBM (n=79) of study EGAS00001003672 using customized gene signatures indicating “Signaling” processes, cellular subsets and stages of microglia and astrocyte and immune cell subsets. ssGSEA demonstrated distinct separation of MBM with high, median or low immunescore regarding expression levels of signature genes, BMCs served as controls. ssGSEA uncovered differentially activated pathways and processes such as MET and STAT3 and interferon signaling, senescence (SenMayo), stress response and tumor inflammation in tumors enriched for reactive microglia and astrocytes and innate and acquired immune cells subsets. b.) Boxplot showing expression levels of HGF in indicated immune cell subsets obtained from DICE. c.-e.) Expression of MET receptor pathway genes in MBM of study EGAS00001003672 showing high or low enrichment of microglia, as determined by levels of microglia-specific genes (SYK, HCK, AIF1/Iba1= microglia score suggests a significant correlation of microglia infiltration and activation of MET receptor signaling. HGF, hepatocyte growth factor (p=1.8e-09); PIK3CG, Phosphatidylinositol-4,5-Bisphosphate 3-Kinase Catalytic Subunit Gamma (p<2.2e-16); PTK2B, Protein Tyrosine Kinase 2 Beta (p<2.2e-16); STAT3, Signal Transducer And Activator Of Transcription 3 (p=5.3e-11); MAP4K1; Mitogen-Activated Protein Kinase Kinase Kinase Kinase 1 (p=3.4e-12). Box and whisker plots show median (center line), the upper and lower quartiles (the box), and the range of the data (the whiskers), including outliers (b-e).

**Supplementary figure 5:** MET-FISH analysis revealed absence of MET receptor amplifications in MBM. a.) Hematoxylin and eosin (H&E) staining shows tumor cell histology (upper row). Fluorescence in-situ hybridization with a MET-specific probe (red) revealed no specific amplifications of the MET gene in MBM (n=7) as compared with centromere control (green) and irrespective of the BRAF/NRAS mutation status. DAPI served as nuclear dye, bars indicate 50 µm. b.) Quantitative representation of FISH analysis, indicating the number of MET copies per nucleus and ratio of MET and CEP7 (Centromer 7). c.) IHC for Iba1 (red) and pMET^Y12^^34/1235^ (brown) revealed absence of activated MET in Iba1^high^ microglia residing in adjacent tissue (upper panel) and MET activation in tumor cells without neighboring TAMs. d.) HGF expression in brain cells residing within the different lobes (FL, frontal; PL, parietal; TL, temporal; OL, occipital lobe) and pons as retrieved from the Allan Brain Atlas (https://portal.brain-map.org/). Box and whisker plots show median (center line), the upper and lower quartiles (the box), and the range of the data (the whiskers), including outliers (d).

**Supplementary figure 6:** Expression of interferon-related genes is enriched in MBM of ITGB7^high^/IScore^high^ phenotype. a.) Heat map indicating expression levels and subset-association of interferon-inducible genes, mediators of interferon signaling and relevant immune cell-expressed markers such as CD3E, CD4, CD8A in MBM (n=79) of study EGAS00001003672. Molecular subsets, category of genes and strength of expression are color coded. b.-d.) Dot plots indicating the significant correlation of *ITGB7*, *IRF1* (Interferon Regulatory Factor 1), *IRF8* and *IFNG* in MBM and EM of the aforementioned study. e.) Dot plots indicating the significant correlation of expression of *HGF*, *IRF1* and *IRF8* in MBM and EM of the aforementioned study. f.-g.) Investigation of expression data of murine BV2 microglia cells (study GSE132739) following interferon (1 U/mL IFNγ, 24h) or control treatment revealed interferon-responsible genes. Interferon treatment significantly increased levels of Itgb7 (p=2.9e-03), Hgf (p=4.4e-02), Mx1 (p=1.2e-02), Cd274 (p=3.6e-02), Irf1 (p=3.1e-02), Cxcl9 (p=4.0e-03) and Aif1 (p=4.0e-03). However, Susd3 was significantly downregulated upon interferon treatment (p=4.0e-02). h.) ssGSEA analysis of MBM with defined signatures showing enrichment of interferon-related signaling among other indicated processes. Box and whisker plots show median (center line), the upper and lower quartiles (the box), and the range of the data (the whiskers), including outliers (f, g).

**Supplementary figure 7:** The mTOR/pS6 signaling is activated in MBM. a.) IHC of MBM (Pat 14) for levels of activated/phosphorylated MET receptor (pMET^Y12^^34/1235^) and mTOR/pS6 (pS6^S2^^35/236^) signaling revealed co-occurrence of both. b.) Co-occurrence of pS6^S235/236^ and MITF. Bars indicate 50 µm. c.) Confocal microscopy imaging of BMC1-M1 and BMC53 cells for levels of NGFR and MET showing a mutually exclusive expression pattern or low level of MET in NGFR^+^ cells. d.) Live cell imaging tracked dose-response of BMC1-M1 cells to increasing doses of ARQ197.

## References

1 Redmer, T. Deciphering mechanisms of brain metastasis in melanoma - the gist of the matter. Mol Cancer 17, 106, doi:10.1186/s12943-018-0854-5 (2018).

2 Srinivasan, E. S., Deshpande, K., Neman, J., Winkler, F. & Khasraw, M. The microenvironment of brain metastases from solid tumors. Neurooncol Adv 3, v121–v132, doi:10.1093/noajnl/vdab121 (2021).

3 Holt, M. G. Astrocyte heterogeneity and interactions with local neural circuits. Essays Biochem 67, 93–106, doi:10.1042/ebc20220136 (2023).

4 Hilscher, M. M. et al. Spatial and temporal heterogeneity in the lineage progression of fine oligodendrocyte subtypes. BMC Biology 20, 122, doi:10.1186/s12915-022-01325-z (2022).

5 Li, Y. et al. Decoding the temporal and regional specification of microglia in the developing human brain. Cell stem cell 29, 620–634 e626, doi:10.1016/j.stem.2022.02.004 (2022).

6 Tan, Y.-L., Yuan, Y. & Tian, L. Microglial regional heterogeneity and its role in the brain. Molecular Psychiatry 25, 351–367, doi:10.1038/s41380-019-0609-8 (2020).

7 Liddelow, S. A. & Barres, B. A. Reactive Astrocytes: Production, Function, and Therapeutic Potential. Immunity 46, 957–967, doi:10.1016/j.immuni.2017.06.006 (2017).

8 Bennett, M. L. & Viaene, A. N. What are activated and reactive glia and what is their role in neurodegeneration? Neurobiol Dis 148, 105172, doi:10.1016/j.nbd.2020.105172 (2021).

9 Tan, Y. L., Yuan, Y. & Tian, L. Microglial regional heterogeneity and its role in the brain. Mol Psychiatry 25, 351–367, doi:10.1038/s41380-019-0609-8 (2020).

10 Mathys, H. et al. Temporal Tracking of Microglia Activation in Neurodegeneration at Single-Cell Resolution. Cell Rep 21, 366–380, doi:10.1016/j.celrep.2017.09.039 (2017).

11 Schwartz, H. et al. Incipient Melanoma Brain Metastases Instigate Astrogliosis and Neuroinflammation. Cancer Res 76, 4359–4371, doi:10.1158/0008-5472.CAN-16-0485 (2016).

12 Colombo, E. & Farina, C. Astrocytes: Key Regulators of Neuroinflammation. Trends Immunol 37, 608–620, doi:10.1016/j.it.2016.06.006 (2016).

13 Rehman, R. et al. Met/HGFR triggers detrimental reactive microglia in TBI. Cell Rep 41, 111867, doi:10.1016/j.celrep.2022.111867 (2022).

14 Yamagata, T. et al. Hepatocyte growth factor specifically expressed in microglia activated Ras in the neurons, similar to the action of neurotrophic factors. Biochemical and biophysical research communications 210, 231–237, doi:10.1006/bbrc.1995.1651 (1995).

15 Nicoleau, C. et al. Endogenous Hepatocyte Growth Factor Is a Niche Signal for Subventricular Zone Neural Stem Cell Amplification and Self-Renewal. Stem Cells 27, 408–419, doi:10.1634/stemcells.2008-0226 (2009).

16 Schetters, S. T. T., Gomez-Nicola, D., Garcia-Vallejo, J. J. & Van Kooyk, Y. Neuroinflammation: Microglia and T Cells Get Ready to Tango. Front Immunol 8, 1905, doi:10.3389/fimmu.2017.01905 (2017).

17 Colonna, M. & Butovsky, O. Microglia Function in the Central Nervous System During Health and Neurodegeneration. Annu Rev Immunol 35, 441–468, doi:10.1146/annurev-immunol-051116-052358 (2017).

18 Caffarel, M. M. & Braza, M. S. Microglia and metastases to the central nervous system: victim, ravager, or something else? J Exp Clin Cancer Res 41, 327, doi:10.1186/s13046-022-02535-7 (2022).

19 Wang, G. et al. Tumor-associated microglia and macrophages in glioblastoma: From basic insights to therapeutic opportunities. Front Immunol 13, 964898, doi:10.3389/fimmu.2022.964898 (2022).

20 Blitz, S. E. et al. Tumor-Associated Macrophages/Microglia in Glioblastoma Oncolytic Virotherapy: A Double-Edged Sword. Int J Mol Sci 23, doi:10.3390/ijms23031808 (2022).

21 Urbantat, R. M. et al. Tumor-Associated Microglia/Macrophages as a Predictor for Survival in Glioblastoma and Temozolomide-Induced Changes in CXCR2 Signaling with New Resistance Overcoming Strategy by Combination Therapy. Int J Mol Sci 22, doi:10.3390/ijms222011180 (2021).

22 Andersen, R. S., Anand, A., Harwood, D. S. L. & Kristensen, B. W. Tumor-Associated Microglia and Macrophages in the Glioblastoma Microenvironment and Their Implications for Therapy. Cancers (Basel) 13, doi:10.3390/cancers13174255 (2021).

23 Radke, J. et al. Decoding molecular programs in melanoma brain metastases. Nature Communications 13, 7304, doi:10.1038/s41467-022-34899-x (2022).

24 Köhler, C. Allograft inflammatory factor-1/Ionized calcium-binding adapter molecule 1 is specifically expressed by most subpopulations of macrophages and spermatids in testis. Cell and Tissue Research 330, 291–302, doi:10.1007/s00441-007-0474-7 (2007).

25 Yoshihara, K. et al. Inferring tumour purity and stromal and immune cell admixture from expression data. Nat Commun 4, 2612, doi:10.1038/ncomms3612 (2013).

26 Ros-Martinez, S., Navas-Carrillo, D., Alonso-Romero, J. L. & Orenes-Pinero, E. Immunoscore: a novel prognostic tool. Association with clinical outcome, response to treatment and survival in several malignancies. Crit Rev Clin Lab Sci 57, 432–443, doi:10.1080/10408363.2020.1729692 (2020).

27 Zhang, Y. et al. Integrin beta7 Inhibits Colorectal Cancer Pathogenesis via Maintaining Antitumor Immunity. Cancer Immunol Res 9, 967–980, doi:10.1158/2326-6066.CIR-20-0879 (2021).

28 Ge, Q. et al. Immunological Role and Prognostic Value of APBB1IP in Pan-Cancer Analysis. J Cancer 12, 595–610, doi:10.7150/jca.50785 (2021).

29 Geirsdottir, L. et al. Cross-Species Single-Cell Analysis Reveals Divergence of the Primate Microglia Program. Cell 179, 1609–1622 e1616, doi:10.1016/j.cell.2019.11.010 (2019).

30 Lafuente, E. M., et al. RIAM, an Ena/VASP and Profilin ligand, interacts with Rap1-GTP and mediates Rap1-induced adhesion. Developmental cell 7, 585–595, doi:10.1016/j.devcel.2004.07.021 (2004).

31 Inagaki, T. et al. The retinoic acid-responsive proline-rich protein is identified in promyeloleukemic HL-60 cells. J Biol Chem 278, 51685–51692, doi:10.1074/jbc.M308016200 (2003).

32 Hoffmann, F. et al. Prognostic and predictive value of PD-L2 DNA methylation and mRNA expression in melanoma. Clin Epigenetics 12, 94, doi:10.1186/s13148-020-00883-9 (2020).

33 Chen, S. et al. Integrin alphaEbeta7(+) T cells direct intestinal stem cell fate decisions via adhesion signaling. Cell research 31, 1291–1307, doi:10.1038/s41422-021-00561-2 (2021).

34 Zhang, L. et al. Single-Cell Analyses Inform Mechanisms of Myeloid-Targeted Therapies in Colon Cancer. Cell 181, 442–459 e429, doi:10.1016/j.cell.2020.03.048 (2020).

35 Schmiedel, B. J. et al. Single-cell eQTL analysis of activated T cell subsets reveals activation and cell type-dependent effects of disease-risk variants. Sci Immunol 7, eabm2508, doi:10.1126/sciimmunol.abm2508 (2022).

36 Weiss, S. A. et al. Melanoma brain metastases have lower T-cell content and microvessel density compared to matched extracranial metastases. J Neurooncol 152, 15–25, doi:10.1007/s11060-020-03619-0 (2021).

37 Griss, J. et al. B cells sustain inflammation and predict response to immune checkpoint blockade in human melanoma. Nat Commun 10, 4186, doi:10.1038/s41467-019-12160-2 (2019).

38 Huang, L. et al. Correlation of tumor-infiltrating immune cells of melanoma with overall survival by immunogenomic analysis. Cancer medicine 9, 8444–8456, doi:10.1002/cam4.3466 (2020).

39 Saul, D. et al. A new gene set identifies senescent cells and predicts senescence-associated pathways across tissues. Nat Commun 13, 4827, doi:10.1038/s41467-022-32552-1 (2022).

40 Pais Ferreira D., et al. Central memory CD8(+) T cells derive from stem-like Tcf7(hi) effector cells in the absence of cytotoxic differentiation. Immunity 53, 985–1000 e1011, doi:10.1016/j.immuni.2020.09.005 (2020).

41 Sade-Feldman, M. et al. Defining T Cell States Associated with Response to Checkpoint Immunotherapy in Melanoma. Cell 176, 404, doi:10.1016/j.cell.2018.12.034 (2019).

42 Connolly, K. A. et al. A reservoir of stem-like CD8(+) T cells in the tumor-draining lymph node preserves the ongoing antitumor immune response. Sci Immunol 6, eabg7836, doi:10.1126/sciimmunol.abg7836 (2021).

43 Fischer, G. M. et al. Molecular Profiling Reveals Unique Immune and Metabolic Features of Melanoma Brain Metastases. Cancer Discov 9, 628–645, doi:10.1158/2159-8290.CD-18-1489 (2019).

44 Desole, C. et al. HGF and MET: From Brain Development to Neurological Disorders. Front Cell Dev Biol 9, 683609, doi:10.3389/fcell.2021.683609 (2021).

45 Zhang, Y. et al. Purification and Characterization of Progenitor and Mature Human Astrocytes Reveals Transcriptional and Functional Differences with Mouse. Neuron 89, 37–53, doi:10.1016/j.neuron.2015.11.013 (2016).

46 Zhang, Y., Jain, R. K. & Zhu, M. Recent Progress and Advances in HGF/MET-Targeted Therapeutic Agents for Cancer Treatment. Biomedicines 3, 149–181, doi:10.3390/biomedicines3010149 (2015).

47 Ramani, N. S., Morani, A. C. & Zhang, S. MET Gene High Copy Number (Amplification/Polysomy) Identified in Melanoma for Potential Targeted Therapy. Am J Clin Pathol 157, 502–505, doi:10.1093/ajcp/aqab171 (2022).

48 Rozeman, E. A. et al. Survival and biomarker analyses from the OpACIN-neo and OpACIN neoadjuvant immunotherapy trials in stage III melanoma. Nat Med 27, 256–263, doi:10.1038/s41591-020-01211-7 (2021).

49 Reijers, I. L. M. et al. The interferon-gamma (IFN-y) signature from baseline tumor material predicts pathologic response after neoadjuvant ipilimumab (IPI) + nivolumab (NIVO) in stage III melanoma. Journal of Clinical Oncology 40, 9539–9539, doi:10.1200/JCO.2022.40.16_suppl.9539 (2022).

50 Subramanian, A. et al. Gene set enrichment analysis: a knowledge-based approach for interpreting genome-wide expression profiles. Proc Natl Acad Sci U S A 102, 15545–15550, doi:10.1073/pnas.0506580102 (2005).

51 Gonzalez, H. et al. Cellular architecture of human brain metastases. Cell 185, 729–745 e720, doi:10.1016/j.cell.2021.12.043 (2022).

52 Dufner, A., Andjelkovic, M., Burgering, B. M., Hemmings, B. A. & Thomas, G. Protein kinase B localization and activation differentially affect S6 kinase 1 activity and eukaryotic translation initiation factor 4E-binding protein 1 phosphorylation. Mol Cell Biol 19, 4525–4534, doi:10.1128/MCB.19.6.4525 (1999).

53 Seip, K. et al. Fibroblast-induced switching to the mesenchymal-like phenotype and PI3K/mTOR signaling protects melanoma cells from BRAF inhibitors. Oncotarget 7, 19997–20015, doi:10.18632/oncotarget.7671 (2016).

54 Yan, Y. et al. Vemurafenib and Cobimetinib Potently Inhibit Ps6 Signaling in Brafv600 Mutation&#x2013;Positive Locally Advanced or Metastatic Melanoma from Brim7 Study. Annals of Oncology 25, iv378, doi:10.1093/annonc/mdu344.9 (2014).

55 Zhao, S. et al. Selective Inhibitor of the c-Met Receptor Tyrosine Kinase in Advanced Hepatocellular Carcinoma: No Beneficial Effect With the Use of Tivantinib? Front Immunol 12, 731527, doi:10.3389/fimmu.2021.731527 (2021).

56 Quail, D. F. & Joyce, J. A. The Microenvironmental Landscape of Brain Tumors. Cancer cell 31, 326–341, doi:10.1016/j.ccell.2017.02.009 (2017).

57 Roesler, R., Dini, S. A. & Isolan, G. R. Neuroinflammation and immunoregulation in glioblastoma and brain metastases: Recent developments in imaging approaches. Clin Exp Immunol 206, 314–324, doi:10.1111/cei.13668 (2021).

58 He, X., Guo, Y., Yu, C., Zhang, H. & Wang, S. Epithelial-mesenchymal transition is the main way in which glioma-associated microglia/macrophages promote glioma progression. Front Immunol 14, 1097880, doi:10.3389/fimmu.2023.1097880 (2023).

59 Zhai, H., Heppner, F. L. & Tsirka, S. E. Microglia/macrophages promote glioma progression. Glia 59, 472–485, doi:10.1002/glia.21117 (2011).

60 Sofroniew, M. V. Astrogliosis. Cold Spring Harbor perspectives in biology 7, a020420, doi:10.1101/cshperspect.a020420 (2014).

61 Herrmann, J. E. et al. STAT3 is a critical regulator of astrogliosis and scar formation after spinal cord injury. J Neurosci 28, 7231–7243, doi:10.1523/JNEUROSCI.1709-08.2008 (2008).

62 Priego, N. et al. STAT3 labels a subpopulation of reactive astrocytes required for brain metastasis. Nat Med 24, 1024–1035, doi:10.1038/s41591-018-0044-4 (2018).

63 Maina, F., Hilton, M. C., Ponzetto, C., Davies, A. M. & Klein, R. Met receptor signaling is required for sensory nerve development and HGF promotes axonal growth and survival of sensory neurons. Genes Dev 11, 3341–3350, doi:10.1101/gad.11.24.3341 (1997).

64 Garcia-Diaz, A. et al. Interferon Receptor Signaling Pathways Regulating PD-L1 and PD-L2 Expression. Cell Rep 29, 3766, doi:10.1016/j.celrep.2019.11.113 (2019).

65 Verhelst, J., Parthoens, E., Schepens, B., Fiers, W. & Saelens, X. Interferon-inducible protein Mx1 inhibits influenza virus by interfering with functional viral ribonucleoprotein complex assembly. J Virol 86, 13445–13455, doi:10.1128/JVI.01682-12 (2012).

66 Acker, G. et al. The CXCR2/CXCL2 signalling pathway - An alternative therapeutic approach in high-grade glioma. Eur J Cancer 126, 106–115, doi:10.1016/j.ejca.2019.12.005 (2020).

67 Jiang, H., Lei, R., Ding, S. W. & Zhu, S. Skewer: a fast and accurate adapter trimmer for next-generation sequencing paired-end reads. BMC Bioinformatics 15, 182, doi:10.1186/1471-2105-15-182 (2014).

68 Love, M. I., Huber, W. & Anders, S. Moderated estimation of fold change and dispersion for RNA-seq data with DESeq2. Genome Biol 15, 550, doi:10.1186/s13059-014-0550-8 (2014).

69 Mootha, V. K. et al. PGC-1alpha-responsive genes involved in oxidative phosphorylation are coordinately downregulated in human diabetes. Nat Genet 34, 267–273, doi:10.1038/ng1180 (2003).

70 Hanzelmann, S., Castelo, R. & Guinney, J. GSVA: gene set variation analysis for microarray and RNA-seq data. BMC Bioinformatics 14, 7, doi:10.1186/1471-2105-14-7 (2013).

71 Biermann, J. et al. Dissecting the treatment-naive ecosystem of human melanoma brain metastasis. Cell 185, 2591–2608 e2530, doi:10.1016/j.cell.2022.06.007 (2022).

72 Schildhaus, H. U. et al. MET amplification status in therapy-naive adeno- and squamous cell carcinomas of the lung. Clin Cancer Res 21, 907–915, doi:10.1158/1078-0432.CCR-14-0450 (2015).

73 Livak, K. J. & Schmittgen, T. D. Analysis of relative gene expression data using real-time quantitative PCR and the 2(-Delta Delta C(T)) Method. Methods 25, 402–408, doi:10.1006/meth.2001.1262 (2001).

