## Supplementary figures and images for "MET receptor activation by stromal cells serves as promising target in melanoma brain metastases"

### Supplementary figure 1: Iba1/AIF1 expression separates MBM

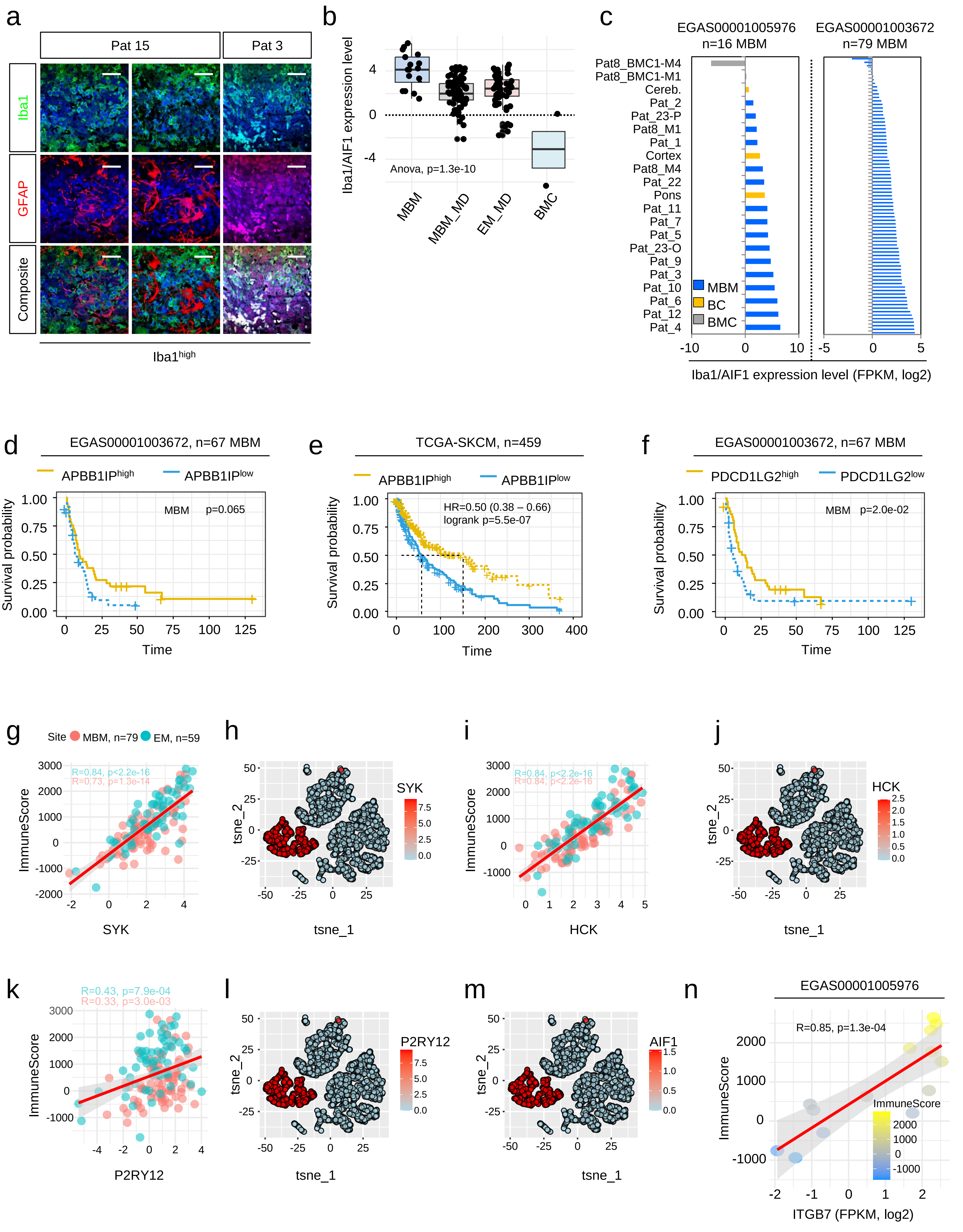


Redmer et al., Supplementary figure 1

### Supplementary figure 2: The indicators of favorable disease course ITGB7, SUSD3 and APBB1IP are broadly expressed among immune cell subsets

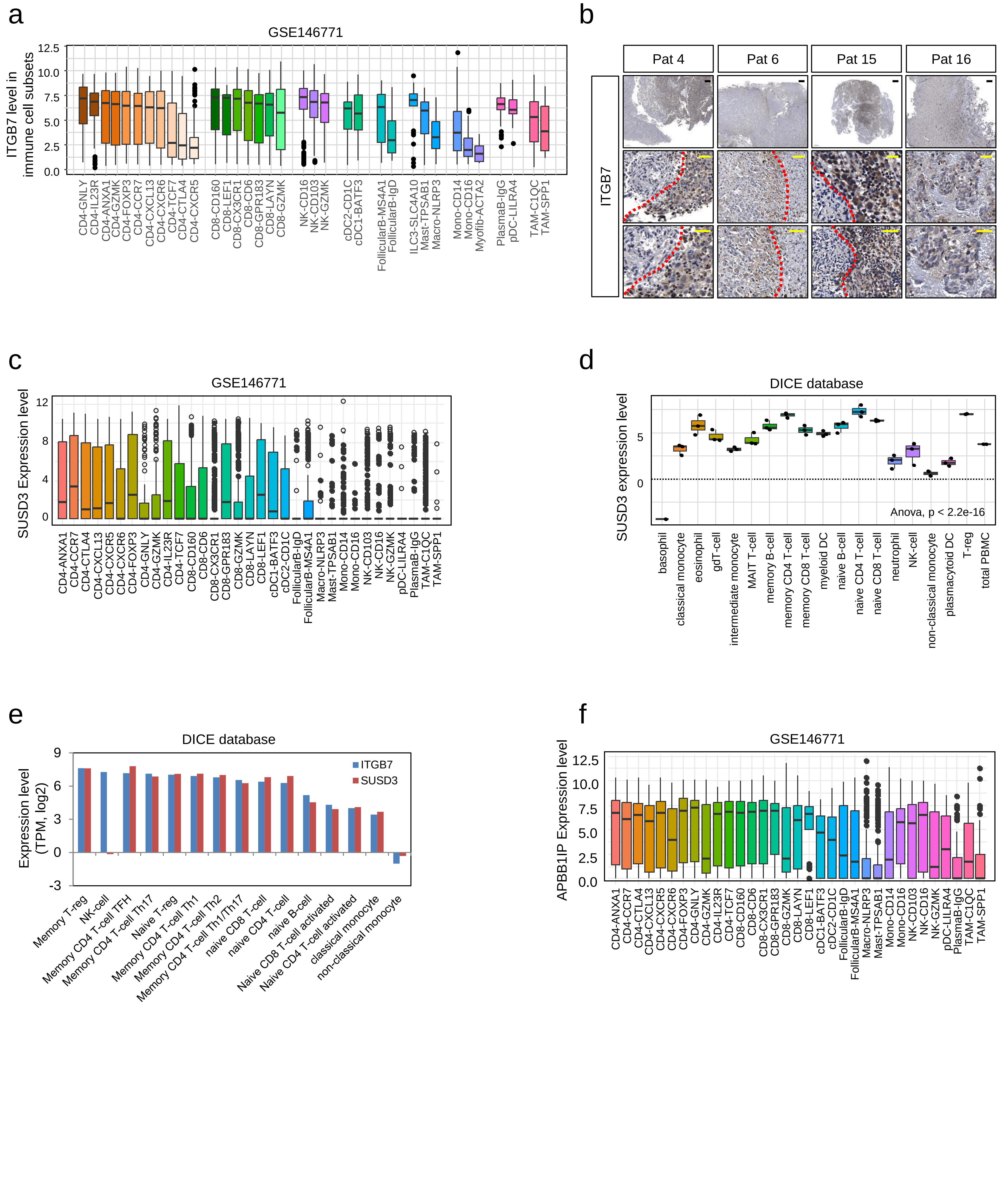


Redmer et al., Supplementary figure 2

### Supplementary figure 3: Methylome profiling uncovered epigenetic regulatory sites in the ITGB7 gene

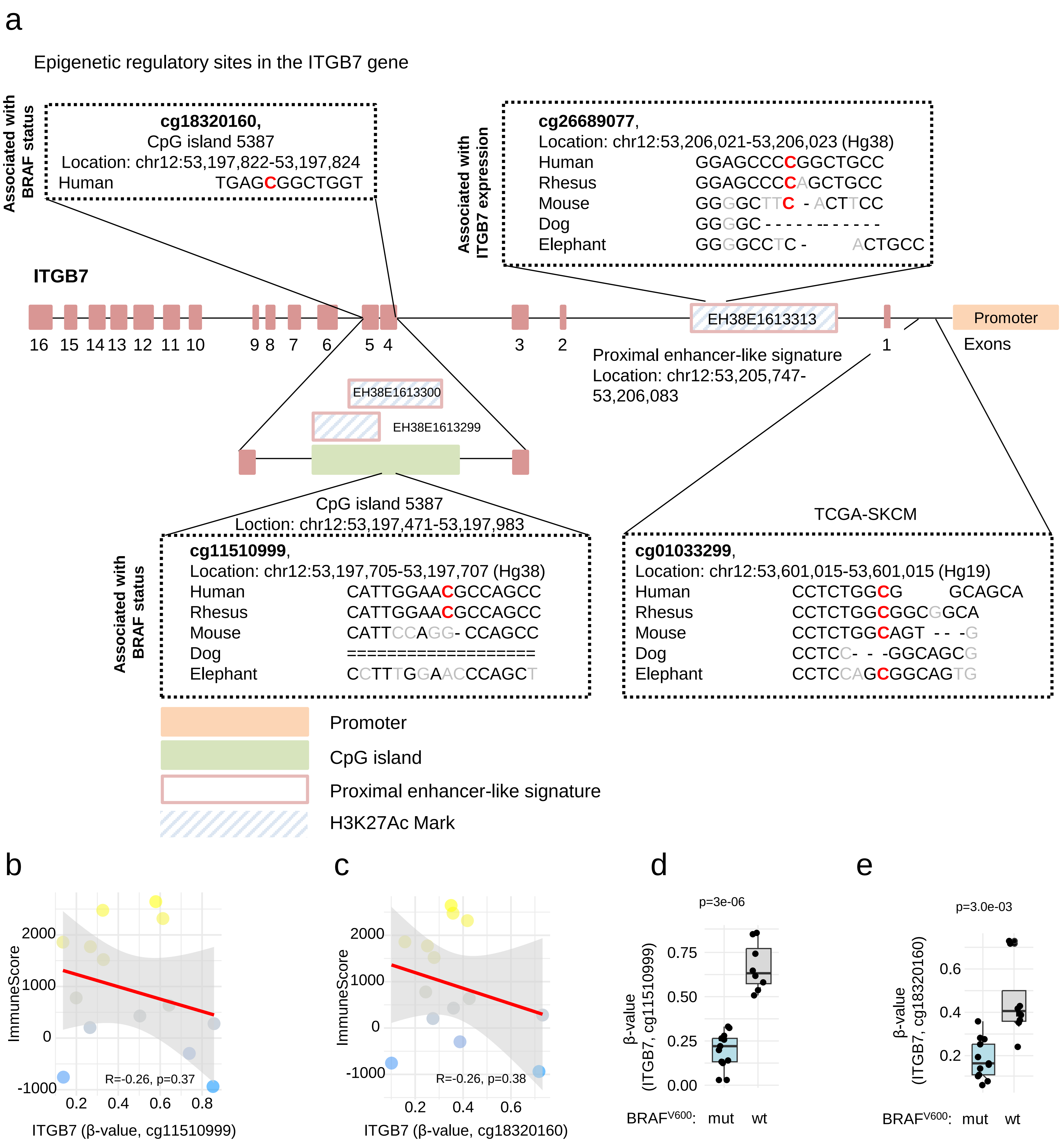


Redmer et al., Supplementary figure 3

### Supplementary figure 4: Single sample GSEA revealed classification of immune molecular subtypes of MBM

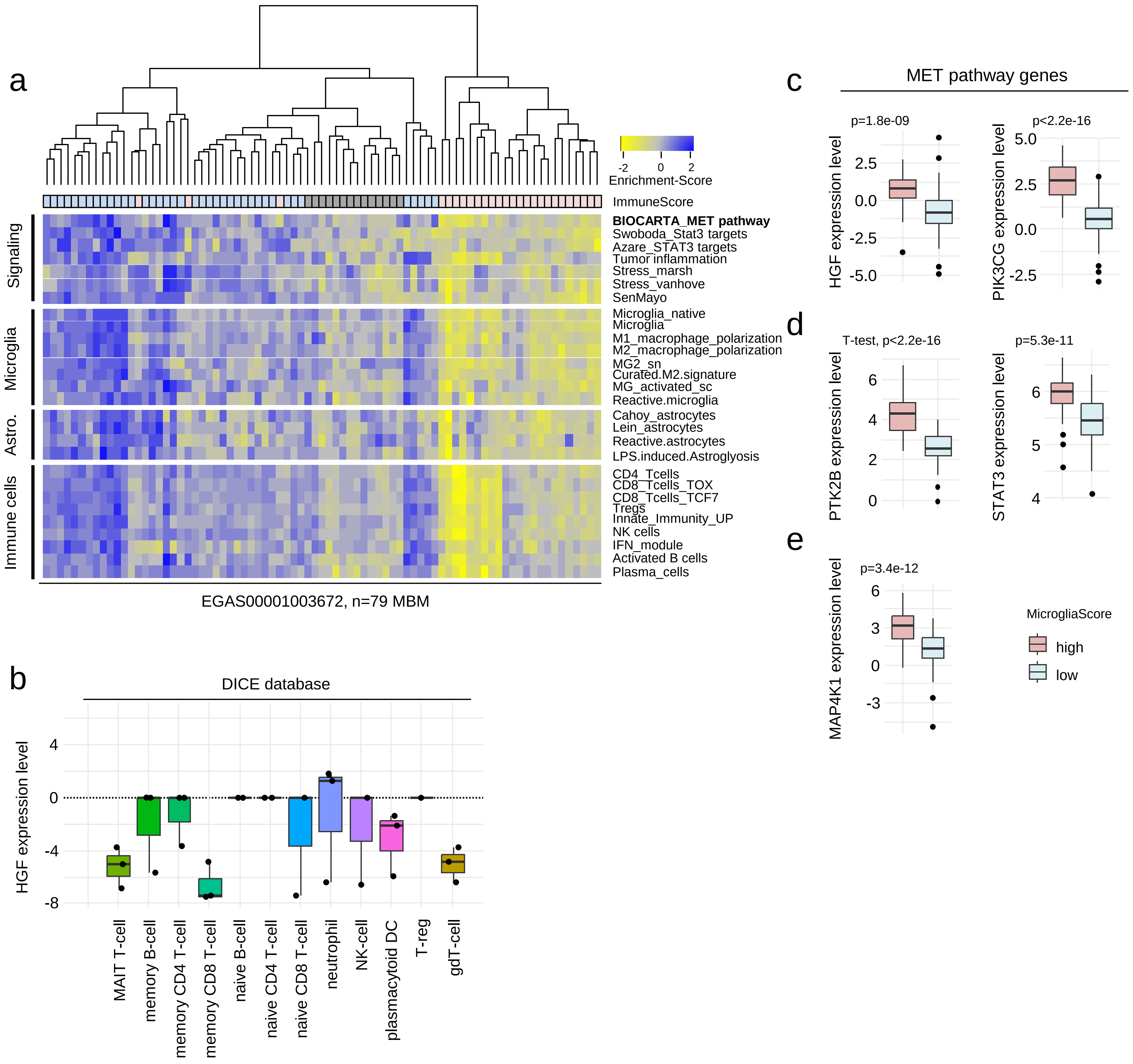


Redmer et al., Supplementary figure 4

### Supplementary figure 5: MET-FISH analysis revealed absence of MET receptor amplifications in MBM

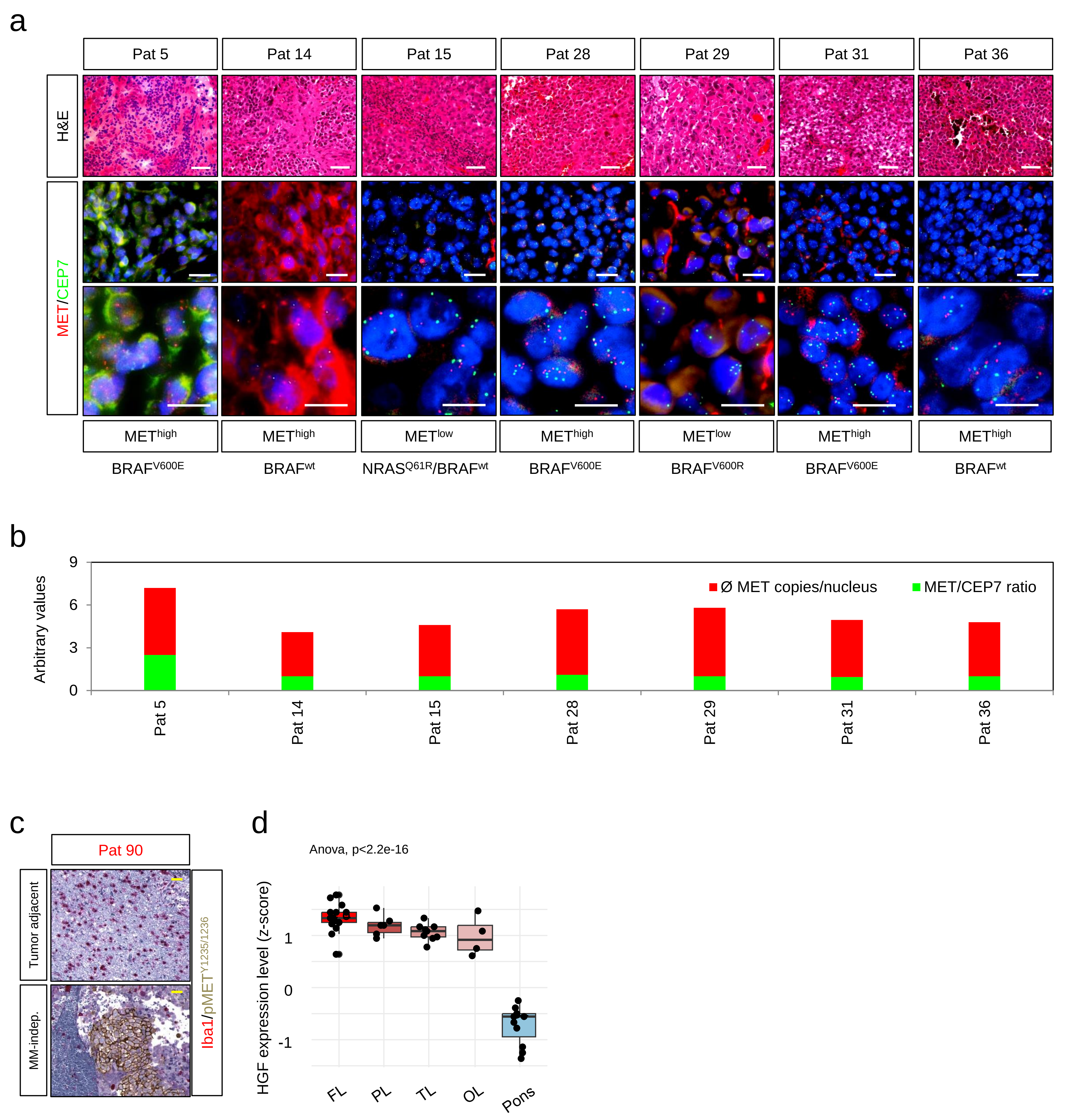


Redmer et al., Supplementary figure 5

### Supplementary figure 6: Expression of interferon-related genes is enriched in MBM of ITGB7high/IScorehigh phenotype

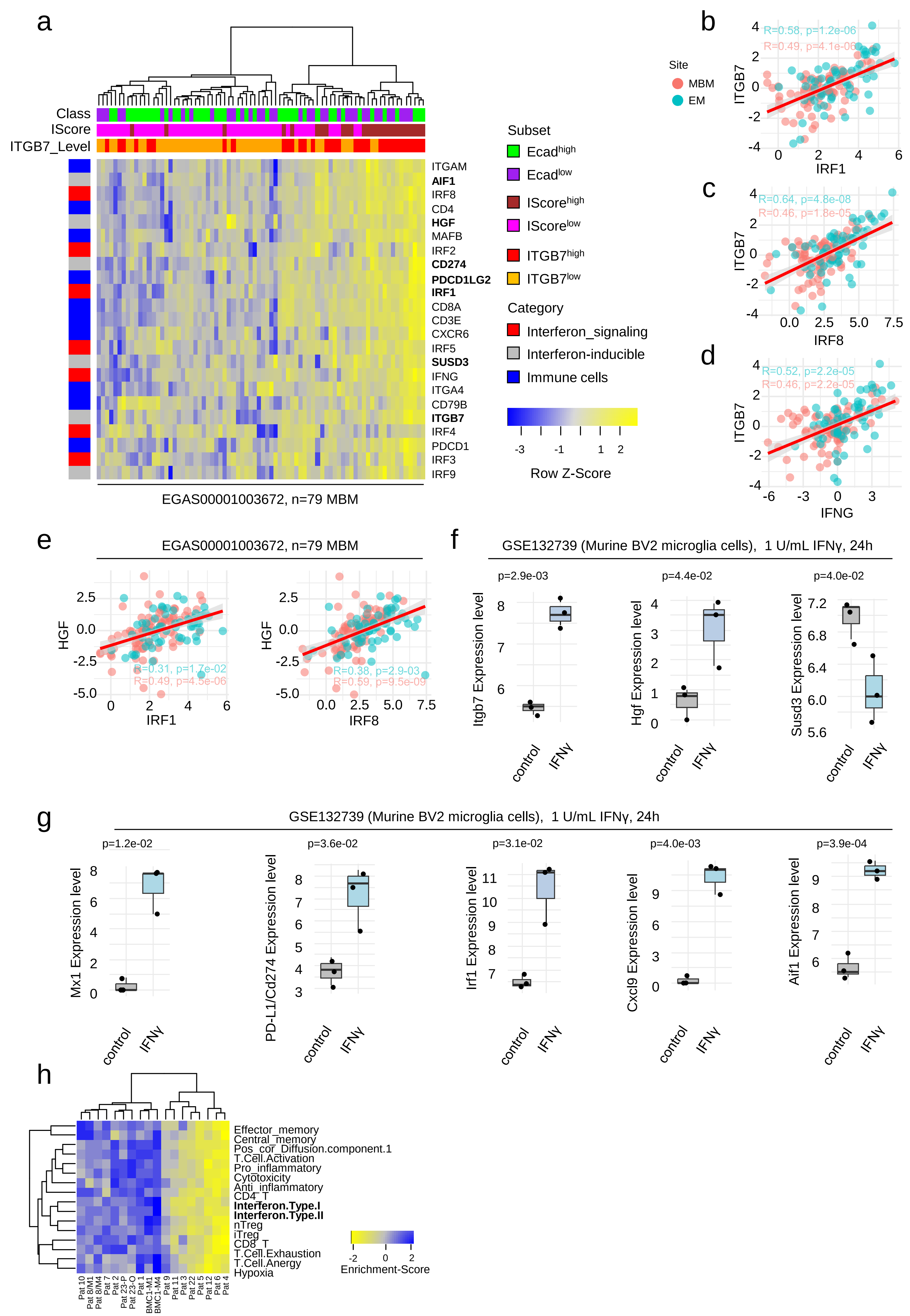


Redmer et al., Supplementary figure 6

### Supplementary figure 7: The mTOR/pS6 signaling is activated in MBM

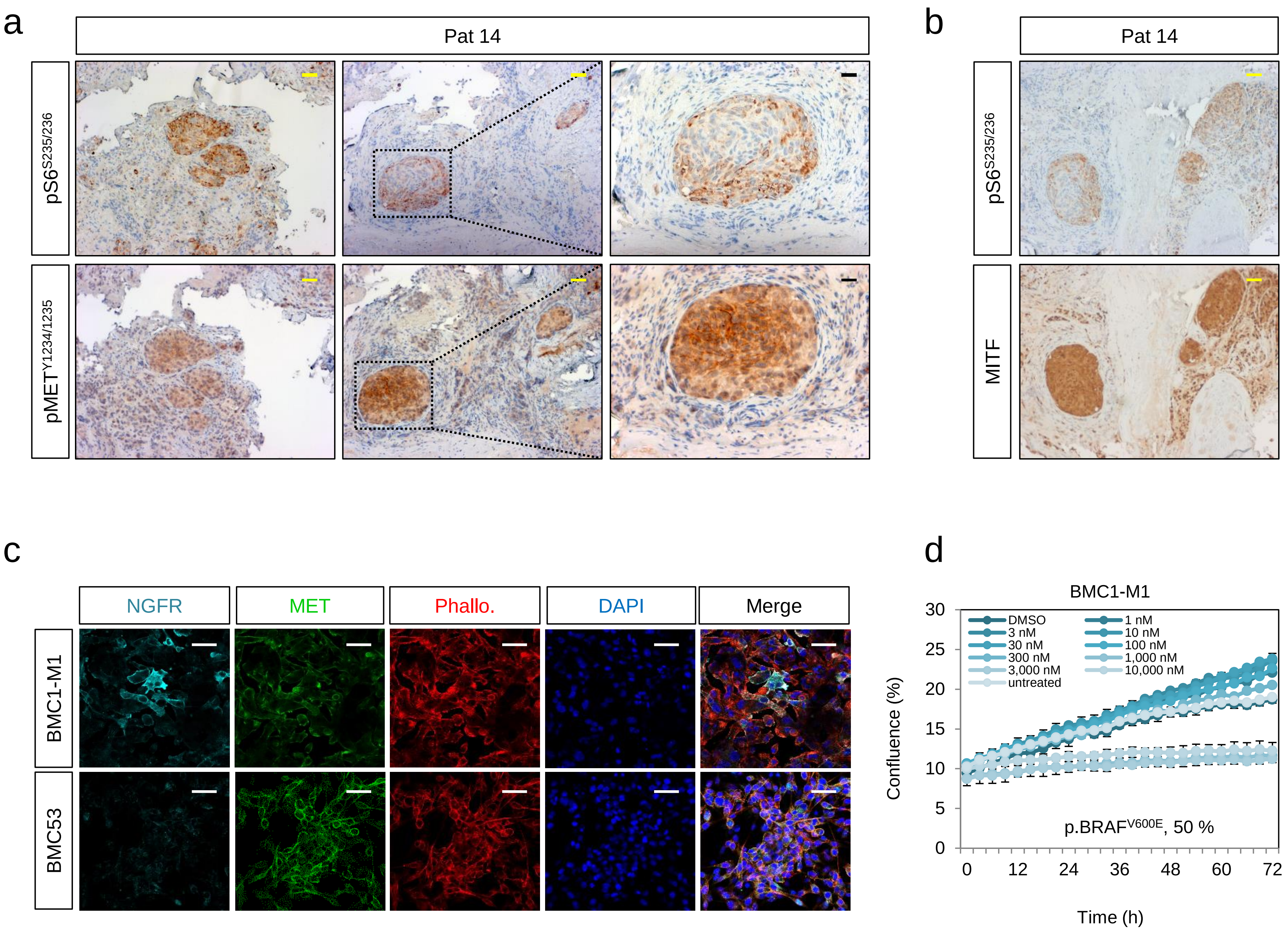


Redmer et al., Supplementary figure 7
